# Gut microbiome alterations precede graft rejection in kidney transplantation patients

**DOI:** 10.1101/2024.05.21.24307660

**Authors:** Johannes Holle, Rosa Reitmeir, Felix Behrens, Dharmesh Singh, Daniela Schindler, Olena Potapenko, Victoria McParland, Harithaa Anandakumar, Nele Kanzelmeyer, Claudia Sommerer, Steffen Hartleif, Joachim Andassy, Uwe Heemann, Michael Neuenhahn, Sofia K. Forslund-Startceva, Markus Gerhard, Jun Oh, Nicola Wilck, Ulrike Löber, Hendrik Bartolomaeus, members of the German Center of Infection Research Transplant Cohort

**Affiliations:** Department of Pediatric Gastroenterology, Nephrology and Metabolic Diseases, Charité – Universitätsmedizin Berlin, Berlin, Germany; Experimental and Clinical Research Center, a cooperation of Charité – Universitätsmedizin Berlin and Max Delbrück Center for Molecular Medicine, Berlin, Germany; German Centre for Cardiovascular Research (DZHK), Partner Site Berlin, Berlin, Germany; Max Delbrück Center for Molecular Medicine in the Helmholtz Association, Berlin, Germany; Department of Preclinical Medicine, Institute for Medical Microbiology, Immunology and Hygiene, Technical University of Munich (TUM), TUM School of Medicine and Health, Munich, Germany; German Center for Infection Research (DZIF), Partner Site München, Germany; German Center for Infection Research (DZIF), Partner Site Braunschweig, Germany; Department of Pediatric Kidney, Liver and Metabolic Diseases, Hannover Medical School, Children’s Hospital, Hannover, Germany; German Center for Infection Research (DZIF), Partner Site Hannover, Germany; Department of Nephrology, University of Heidelberg, Heidelberg, Germany; German Center for Infection Research (DZIF), Partner Site Heidelberg, Germany; Paediatric Gastroenterology and Hepatology, University Children’s Hospital Tübingen, Tübingen, Germany; German Center for Infection Research (DZIF), Partner Site Tübingen, Germany; Klinik für Allgemeine-, Viszeral-, und Transplantationschirurgie, Klinikum der Universität München, Munich, Germany; Department of Nephrology, Technical University of Munich, Munich, Germany; Department of Pediatric Nephrology, University Children’s Hospital, University Medical Center Hamburg-Eppendorf, Hamburg, Germany; Department of Nephrology and Medical Intensive Care, Charité – Universitätsmedizin Berlin, Berlin, Germany

## Abstract

**Background:** Kidney transplantation (KT) is the optimal treatment for end-stage kidney disease, with graft survival critically affected by the recipient’s immune response. The role of the gut microbiome in modulating this immune response remains underexplored. Our study investigates how microbiome alterations might associate with allograft rejection.

**Methods:** We analyzed existing biomaterials of a multicenter prospective study involving 217 KT recipients and 28 kidney donors from the German Center for Infection Research. Changes in the gut microbiome were analyzed using 16S rRNA gene amplicon sequencing and functional predictions (PICRUSt2) and quantitative PCRs for the production potential of propionate and butyrate. Propensity score matching was utilized to compare patients who experienced graft rejection with those who did not.

**Results:** The gut microbiome showed gradual recovery post-KT, marked by an increase of Shannon diversity and SCFA-producing bacterial taxa. However, prior to graft rejection, significant alterations were noted in microbiome composition, characterized by a decrease in microbial diversity and SCFA-producing taxa. Post-rejection analysis revealed normalization of these microbiome features. Functional analysis highlighted a decreased potential for SCFA production in patients prior to rejection. Comparison to published microbiome signatures from chronic kidney disease (CKD) patients demonstrated a partial overlap of the microbiome alterations preceding graft rejection with the alterations typically found in CKD.

**Conclusions:** Our findings suggest that alterations in the gut microbiome composition and function may precede and influence KT rejection, suggesting potential for use as biomarker and early therapeutic microbiome-targeting interventions to improve transplant outcomes.

**Key points:**

– CKD-related microbiome alterations recover over time after transplantation mirroring CKD-to-health transition
– Microbiome alterations with lowered production potential of short-chain fatty acids precede graft rejection, likely influencing graft immunity
– The persistence of CKD-associated microbiome characteristics in rejection patients opens avenues for innovative treatment strategies.

## 1. Introduction

Kidney transplantation (KT) represents the best treatment option for patients with advanced kidney failure (CKD G5)^1^. Despite ongoing efforts to prevent the progression of CKD and related comorbidities, successful KT and graft survival, especially prevention of graft rejection, are of outmost importance. Graft rejection is the medium- and long-term complication with highest impact on graft and subsequently recipient survival^2^. Whether a patient experiences acute or chronic graft rejection or not is, according to the current scientific understanding, largely dependent on immune mechanisms, the modifiers of which still remain poorly understood^3^.

Of late, the gut microbiome gained considerable attention as key modulator of the immune system in both healthy and disease conditions^4^. Microbiota, their metabolites and associated molecules interact with numerous host organs including mucosa-associated and systemic immune cells and thereby shape host immunity and inflammation^5^. Patients with CKD exhibit marked alterations to their gut microbiome composition and subsequent dysregulation of metabolite abundance^6^. In particular, a switch from saccharolytic to proteolytic fermentation is observed^7^. On one hand this leads to lower intestinal and systemic concentrations of short-chain fatty acids (SCFA)^8^, previously identified as inducers of regulatory immunity^9^. On the other hand, increased levels of pro-inflammatory metabolites, such as tryptophan-derived indoxyl sulfate, are observed^8^.

After KT, CKD-related microbiome alterations can partially persist, presumably by contribution of additional factors such as immunosuppression^10^, and are associated with clinical events like post-transplant diarrhea^11,12^. A recent study rigorously demonstrated an association between lower gut microbial diversity in patients with solid organ transplantation and increased overall mortality^13^. A growing body of evidence implicates the gut microbiota in alloimmunity and graft rejection. Germ-free mice and mice treated with antibiotics show prolonged skin graft survival compared to conventional mice, suggesting that alloimmunity is modulated by microbiota^14^. Furthermore, recent studies indicate that gut microbes directly affect immune-regulatory cells (regulatory T cells, Th17 cells)^15,16^, with SCFA playing a central role in this relationship^17^. Studies in mice showed that SCFA supplementation mediates donor-specific tolerance to kidney allograft through induction of Foxp3^+^ regulatory T-cells^18^. Finally, human interventional studies are underway, aiming to improve CKD-associated microbiome alterations by prebiotic inulin supplementation^19^ or at improving immune regulation by supplementation of regulatory T cells^20^.

In this study, we analyzed longitudinal changes in the composition and function of the gut microbiome of KT recipients enrolled in the transplant cohort (Tx cohort) of the German Center of Infectious Diseases (DZIF)^21^ and connect them with the clinical outcomes, specifically graft survival and rejection. In a propensity score-matched subcohort we identify microbial alterations preceding graft rejections and transplant dysfunction.

## 2. Materials and methods

### Study Population and Design

All patients analyzed in this study were part of the Tx cohort of the DZIF, which is a multicenter prospective cohort study conducted at four German transplant centers (University Hospitals in Heidelberg, Munich (Technical University and Ludwig Maximilian University) and Tübingen)^21^. Together, these centers cover over 20% of solid organ transplants in Germany^22^ providing a representative picture of post-transplant course in Germany.

For the present study, we analyzed data from all kidney allograft recipients who consented to participate and received a kidney transplant between July 2014 and July 2021. Samples were collected between March 2017 and September 2021. Patients receiving multi-organ transplantation or patients who underwent previous solid organ transplantation were excluded (Supplemental Figure 1). Moreover, participants without fecal samples were excluded from our analysis. Ethics approval was obtained from all participating centers (Heidelberg #S-585/2013, TU Munich #5926/13, LMU Munich #380-15, Tübingen #327/2014BO1), and all participants provided written informed consent. The experimental analysis of fecal samples, including metadata analysis, was approved by the ethics board of Charité – Universitätsmedizin Berlin (EA2/208/21).

The study design, including biomaterial collection, was described previously^21^. Fecal samples were collected using DNA stabilizing buffer (STRATEC). Study visits were performed immediately before KT, at months 3, 6, 9, and 12 after KT and when infections or rejection events were detected. Clinical and laboratory data were collected at each study visit.

### Standard of care treatment

The patients received a standard triple-drug combination of immunosuppressants (comprising a calcineurin inhibitor, mycophenolate (MPA), and corticosteroids), initially frequently together with an interleukin 2 receptor antagonist (basiliximab). The prophylaxis and surveillance strategy for infectious complications was suggested to be performed according to KDIGO 2009 guidelines^23^, including antiviral prophylaxis with valganciclovir for patients at high risk for cytomegalovirus (CMV) infections, trimethoprim-sulfamethoxazole prophylaxis against *Pneumocystis jirovecii* and urinary tract infections, *Candida* prophylaxis with oral nystatin or amphotericin B.

### Matching of rejection/non-rejection patients

We conducted our analysis using R (4.3.2), employing MatchIt (4.5.5)^24^ package for sample matching. Patients with histologically proven rejection events were assessed (T cell-mediated rejection (TCMR, Banff category 4) and Borderline (Suspicious) for acute TCMR (Banff category 3))^25^, if the rejection occurred within five years after KT and fecal sampling was obtained before the rejection event (time frame 781 days before the rejection till 3 days after rejection). Patients without sample before rejection and those with-mediated rejection (ABMR) were excluded from the analyses, yielding 33 patients with rejection events. Stool samples taken closest before the rejection event were selected for analyses. Control samples were matched based on the absence of rejection and similar baseline characteristics, using nearest neighbour matching with a 2:1 ratio, matching two controls for each rejection case based on the factors primary condition, donor type, time since transplantation, age, and gender. Of the selected 99 patients, one rejection patient was excluded because of primary graft dysfunction (patient still required dialysis three months after KT) and six patients without rejection were excluded because of sustained poor kidney function (plasma creatinine > 2.5 mg/dL).

Detailed information on clinical data preparation, sample collection and processing, 16S amplicon sequencing, SCFA production gene targeting assay (qPCR) and re-analysis of the CKD dataset^26^ can be found in the online only supplemental material.

### Statistical Analysis

Continuous variables were expressed as medians and interquartile ranges (IQRs). Categorical variables were presented as numbers and percentages. All statistical analyses were conducted using R (4.2.3).

Analysis of alpha diversity and multivariate analysis.

Alpha diversity and multivariate analysis were performed on OTU level on the non-rarefied data. Different metrics for alpha Diversities were calculated using the R-package microbiome (v.1.12.0)^27^. Differences in alpha Diversity were tested using Wilcoxon rank sum test on a significance level of 0.05. For multivariate analysis, Bray-Curtis indices were obtained using the R-package phyloseq (v.1.34.0)^28^ and tested via PERMANOVA using adonis2 of the R-package vegan (v.2.6-4)^29^.

#### Cross-sectional comparison

For the association analysis, the R-package metadecondfoundR (v.0.2.8) was used^30^. The testing was performed on the rarefied data (minimum read count 10270 reads) and all taxonomic levels with a minimum prevalence of 0.1. Wilcoxon rank sum test was performed for binary variables, Spearman‘s correlation for continuous variables, and Kruskal-Wallis test for ordinal variables. The obtained p-values were adjusted using the Benjamini-Hochberg false discovery rate (FDR) correction. In total, 65 clinical variables were considered as confounding factors (Supplemental Table 1). An association was considered statistically significant when FDR < 0.1.

For individual comparisons of clinical and qPCR parameters, Mann-Whitney *U* test was performed and p < 0.05 was considered statistically significant.

#### Longitudinal Analysis

For the longitudinal analysis, LongDat (v.1.1.2)^31^ was used, building negative binomial generalized linear mixed models over time after KT, controlling for sample origin by a random intercept. The models were built on rarefied data with default filtering^31^. FDR was controlled by Benjamini-Hochberg procedure. A change in abundance over time as considered statistically significant when FDR < 0.1.

Diversity parameters during different periods before and after KT were compared using Mann-Whitney *U* test and p < 0.05 was considered statistically significant.

#### Functional capacity prediction using Picrust2

PICRUSt2^32^ obtained abundances of the KEGG orthologs (KOs) were z-score scaled and tested for significance with a linear model correcting for age and sex (Benjamini-Hochberg-corrected FDR < 0.05). All significant KOs were mapped to the corresponding GOmixer modules^33^ and tested for group difference using Wilcoxon rank sum test with a significance level of FDR < 0.05 using the online tools of GOmixer.

## 3. Results

### Gut microbiota composition recovers gradually post-kidney transplantation

We used medical data and fecal samples from the DZIF Transplant Cohort. This multi-center cohort consists of nearly 2400 patients undergoing solid organ transplantation. Here, we focused on fecal samples from patients undergoing KT (n=562 samples, from n= 245 individuals), including kidney donors (n=28) and, if available, pre-transplantation samples (n=26). Baseline characteristics of the cohort can be found in Table 1.

**Table 1.** Clinical baseline characteristics total cohort. Kidney transplant (KT) donors, pre-KT CKD patients and post-KT CKD patients from the transplant cohort of the German Center of Infectious Diseases (DZIF) were analyzed in this study. Data are presented as median and interquartile range or absolute values and percentages as appropriate.

|  | donors | pre-KT | post-KT | N |
| --- | --- | --- | --- | --- |
| individuals (N) | 28 | 26 | 213 |  |
| samples | 28 | 26 | 508 |  |
| samples per participant | 1 | 1 | 2 (2-3) |  |
| age (years) | 53 (46-68) | 52 (34-60) | 54 (40-63) | 28, 26, 213 |
| female | 20 (71%) | 6 (25%) | 70 (33%) | 28, 24, 211 |
| BMI (kg/m<sup>2</sup>) | 26.1 (24.0-28.3) | 25.1 (21.6-27.3) | 24.6 (21.6-28.0) | 22, 26, 210 |
| time since KT (days) | NA | NA | 294 (148-715) | 213 |
| living donor | NA | NA | 81 (38%) | 212 |
| ABO incompatible KT | NA | NA | 18 (9%) | 204 |
| rejection | NA | NA | 76 (35%) | 213 |
| kidney diagnosis |  |  |  | 26, 213 |
| cystic | NA | 4 (15%) | 43 (20%) |  |
| diabetic | NA | 2 (8%) | 14 (7%) |  |
| glomerular | NA | 10 (38%) | 71 (33%) |  |
| hereditary/congenital | NA | 6 (23%) | 25 (12%) |  |
| hypertensive | NA | 0 | 16 (8%) |  |
| other/unknown | NA | 4 (15%) | 37 (17%) |  |
| tubulointerstitial | NA | 0 | 7 (3%) |  |
| primary graft dysfunction | NA | NA | 8 (4%) | 209 |
| delayed graft function | NA | NA | 38 (18%) | 218 |
| in-patient length of stay | NA | NA | 19 (14-26) | 210 |

We analyzed the microbiome composition using 16S rRNA gene amplicon sequencing. For longitudinal analysis post-KT, samples were grouped according to sampling time, namely 0-3 months (n=69), 3-12 months (n=226) 12-24 months (n=94), 24-36 (n=66), and over 36 months (n=53) post-KT (Supplemental Figure 1). Principal Coordinates Analysis (PCoA) of Bray-Curtis dissimilarity revealed an overall stable microbiome composition post-KT, while the healthy donors and pre-KT groups differed (Fig 1A). While the detected number of operational taxonomic units (OTU, a proxy for bacterial taxa) was mostly stable post-transplantation (Fig 1B), we observed significant shifts in the alpha diversity as quantified by Shannon diversity index (Fig 1C). While Shannon diversity decreased before and in the first year after transplantation, it gradually increased again over time post-KT. We observed a similar trend for Simpson evenness, although most comparisons were not significant (Fig 1D).

**Figure 1.**
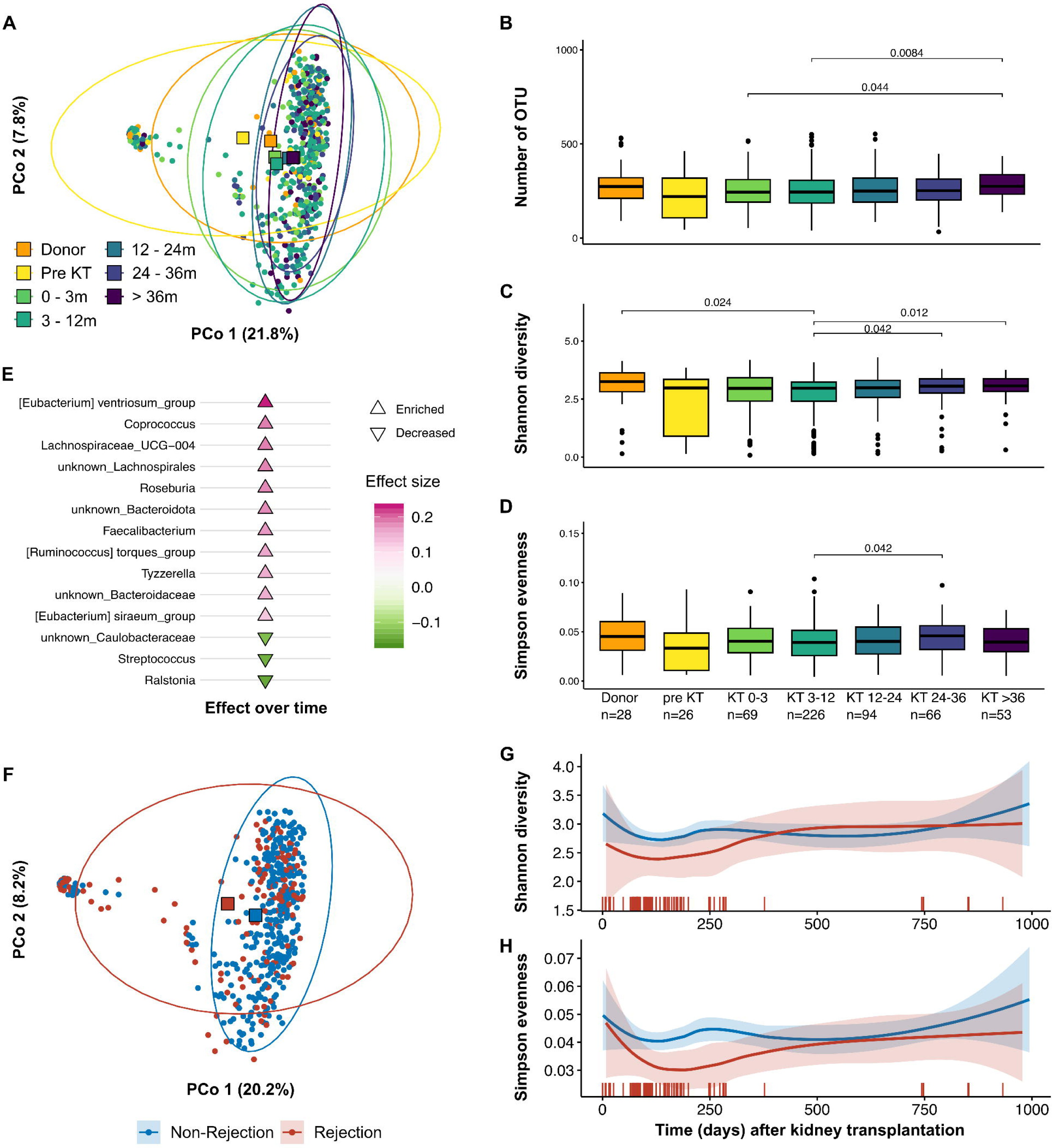
Longitudinal changes to the gut microbiome after kidney transplantation and impact of allograft rejection. 16S rRNA gene amplicon sequencing from fecal material from kidney transplantation (KT)-related samples from the transplant cohort of the German Center of Infectious Diseases (DZIF). Samples were grouped according to healthy kidney donors, pre-KT, 0-3 months post-KT, 3-12 months, 12-24 months and over 24 months. A) PCoA based on Bray-Curtis dissimilarity. Quantification of the B) Number of detected OTU, C) Shannon diversity, and D) Simpson evenness. E) Cuneiform plot displaying significantly altered bacteria post-transplantation over time. Microbial diversity in kidney transplant recipients with and without graft rejection at any point is shown in F-H. F) Principal Coordinates Analysis (PCoA) based on Bray-Curtis dissimilarity illustrates the distinct clustering of patients who experienced graft rejection (red circles) from those without rejection (blue circles). Red and blue squares mark the respective centroids. G+H) Longitudinal analysis of microbial diversity, depicted through Shannon diversity and Simpson evenness indices, shows a reduction in both measures within the first-year post-transplantation for patients undergoing rejection. Vertical red bars indicate individual rejection events over time, coinciding with alterations in microbial diversity.

We performed longitudinal analysis using LongDat^31^ to identify bacterial taxa regulated over time post-KT. Post-transplantation, typical SCFA-producing genera like *Coprococcus*^34^, *Lachnospiraceae*^35^, *Roseburia*^36^, *Faecalibacteria*^37^ and *Ruminocococcus* torques group^38^ increased significantly over time (Fig 1E), suggesting an improvement of CKD-associated microbiome alterations, specifically the impaired production of SCFA as one of its hallmarks^6,8^. Furthermore, we observed a decrease in *Streptococcus*, which was recently linked with subclinical atherosclerosis^39^ (Fig 1E). In summary, our analysis demonstrates a dynamic regeneration of the microbiome over time after KT towards a more physiological state after three years or more post-KT.

### Kidney transplant rejection profoundly impacts microbiome composition

Since we observed dynamic microbiome changes over time in KT patients, we aimed to understand how transplant rejection events influence this process and, vice versa, how microbiome alterations may impact allograft immunity. Therefore, we identified fecal samples (n= 157) from patients (n= 76) with biopsy-proven rejection events at any timepoint after KT (Table 1). PCoA of Bray-Curtis dissimilarity showed a distinct clustering of patients experiencing graft rejection compared to KT patients with no reported rejection (Fig 1F). While the number of detected OTUs was constant over time (Supplemental Figure 2), Shannon diversity and Simpson evenness were reduced during the first year post-transplantation in rejection patients (Fig 1G-H), which was co-incident with the allograft rejection. Lastly, we performed univariate analyses on genus level using metadeconfoundR^30^. Corresponding to the altered composition in the multivariate analysis, we found a large number of differentially abundant genera (Supplemental Figure 3). Interestingly, the majority of observed effects related to rejection showed a contrasting trend in correlation with the duration between sampling and kidney transplant rejection event (Fig S3). This correlation indicates either microbiome alteration preceding the rejection event, changes in the microbiome post-rejection, or both. Of note, our analysis indicates that many effects were confounded by patient age and distance to KT.

Taken together, patients who experience KT rejection have an altered gut microbiome composition. We hypothesize, that these changes might precede transplant rejection, as the alterations associate with the time between sampling and KT rejection event.

### Microbiome alterations precede kidney transplant rejection

To overcome the inherent limitations of the cohort that is unequally distributed age and distance to KT of rejection and non-rejection patients and to further understand which alterations to the microbiome precede KT rejection, we performed propensity score matching of patients with available fecal samples before KT rejection to patients without KT rejection. Matched pre-rejection and non-rejection groups were comparable in age, sex, donor type, underlying CKD disease category and time between KT and sample (Supplemental Figure 4). Baseline characteristics of this subgroup of patients are shown in Table 2.

**Table 2.** Clinical baseline characteristics of the propensity score-matched cohort. Kidney transplant (KT) recipients from the transplant cohort of the German Center of Infectious Diseases (DZIF) were analyzed in this study. Samples were obtained after KT from all patients, but before rejection in the rejection group. Data are presented as median and interquartile range or absolute values and percentages as appropriate. Primary graft dysfunction: patient still required dialysis three months after KT. Delayed graft function: patient required one to three dialysis treatments after KT.

|  | non-rejection | rejection | all | N |
| --- | --- | --- | --- | --- |
| individuals (N) | 60 | 32 | 92 |  |
| age (years) | 55 (44-63) | 58 (47-62) | 56 (45-63) | 92 |
| female | 22 (37%) | 11 (34%) | 33 (36%) | 92 |
| BMI (kg/m<sup>2</sup>) | 23.4 (20.8-27.6) | 24.6 (21.5-27.2) | 24.1 (21.1-27.5) | 92 |
| sample days after KT | 111 (85-175) | 114 (84-192) | 112 (85-181) | 92 |
| rejection days after KT | NA | 173 (132-282) | NA | 92 |
| living donor | 20 (33%) | 10 (31%) | 30 (33%) | 92 |
| ABO incompatible KT | 5 (8%) | 1 (3%) | 6 (7%) | 92 |
| HLA mismatches (HLA-A, HLA-B, HLA-DR) | 2 (2-3) | 3 (2-4) | 3 (2-4) | 79 |
| primary graft dysfunction | 0 | 0 | 0 | 88 |
| delayed graft function | 9 (15%) | 2 (6%) | 11 (12%) | 92 |
| in-patient length of stay (days) | 19 (14-24) | 18 (14-26) | 19 (14-24) | 91 |
| recipient CMV<sup>+</sup> | 26 (46%) | 14 (47%) | 40 (46%) | 87 |
| recipient EBV<sup>+</sup> | 35 (83%) | 18 (90%) | 53 (85%) | 62 |
| bacterial infections before sample | 32 | 20 | 52 | 92 |
| patients with bacterial infection before sample | 17 (28%) | 20 (34%) | 37 (40%) | 92 |
| viral infections before sample | 12 | 23 | 35 | 92 |
| patients with viral infection before sample | 9 (15%) | 14 (44%) | 23 (25%) | 92 |

Rejection patients exhibited impaired renal function as compared to non-rejection patients as displayed by an increase in plasma creatinine within the first year post-KT, which remained the case during follow-up (Fig 2A). This corresponds to most rejections in our cohort occurring within one year post-KT (Table 1 and 2). However, both rejection and normal progress patients reached a similar minimal creatinine as well as time to minimal creatinine (Fig 2B), indicating that initial graft function was comparable between both groups. After the rejection event, kidney transplants showed sustained graft dysfunction as indicated by higher minimal detected creatinine values after their rejection event as well as higher last recorded creatinine (Fig 2B). Of note, HLA mismatch grade, rate of AB0 incompatibility and delayed graft function were comparable between both groups (Table 2).

**Figure 2.**
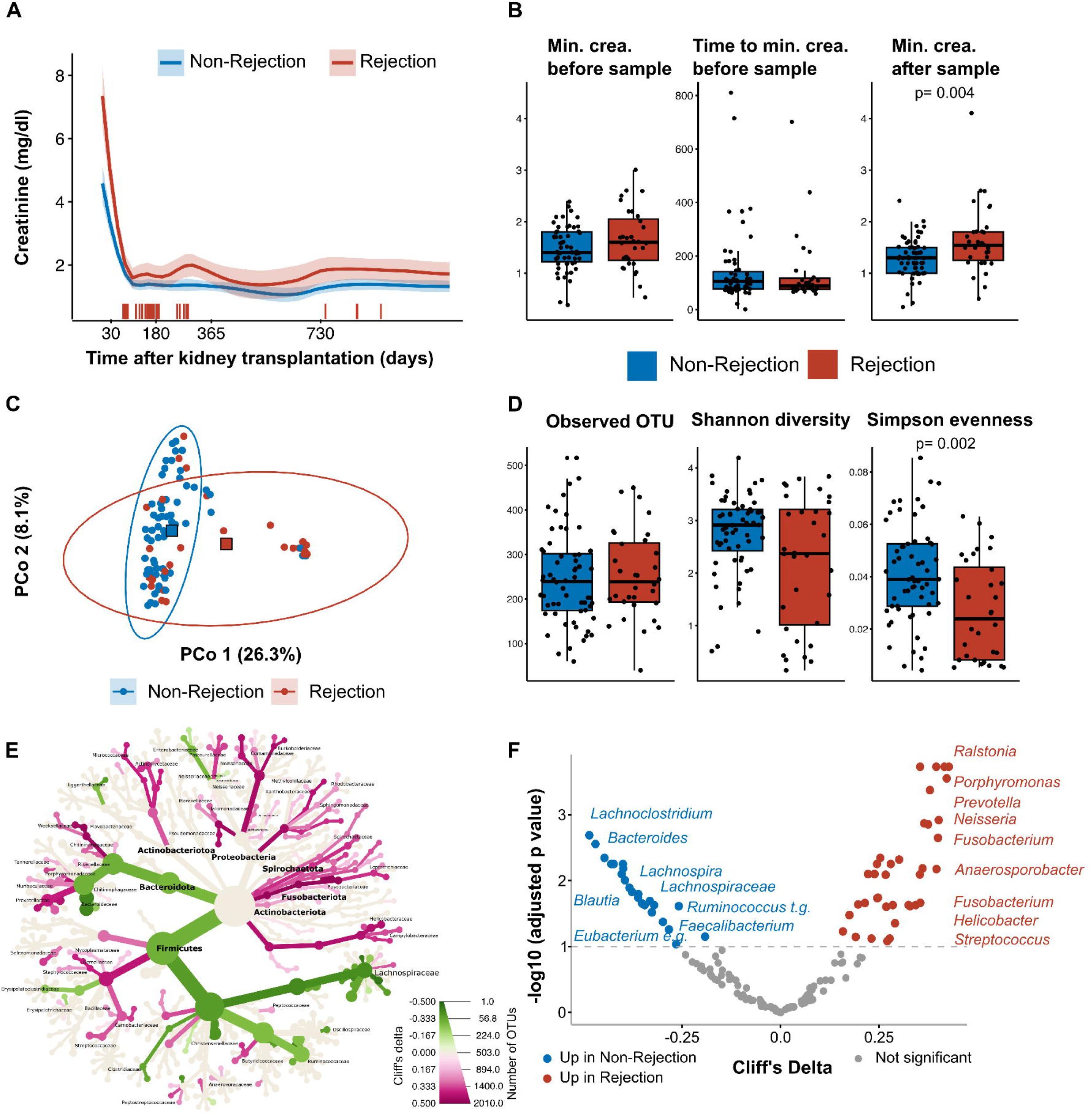
Microbiome alteration precede kidney transplant rejection. Patients with available fecal samples prior to kidney rejection (Rejection group) for propensity score matched (1:2) to patients without kidney rejection (Non-Rejection group). A) Plasma creatinine levels are depicted over time, displaying a consistent increase within the first year post-transplant in patients who experienced graft rejection (red), with these elevated levels persisting at follow-up. Vertical red bars indicate individual rejection events over time. B) Analysis of minimum creatinine levels before and after sampling, along with the time taken to reach the first. C) Gut microbial communities of the two groups based on Bray-Curtis dissimilarity, demonstrate clear separation, with the rejection group (red circles) showing distinct clustering from the non-rejection group (blue circles). Squares represent the centroids of the groups. D) Microbial diversity metrics, including number of detected OTUs, Shannon diversity, and Simpson evenness. E) Differences in microbial taxa between groups, highlighting a decrease in short-chain fatty acid (SCFA)-producing bacteria in the rejection group and an enrichment of genera commonly elevated in chronic kidney disease (CKD) and disease conditions. Bacteria are shown in a phylogenetic tree where green lineages represent bacteria depleted and pink bacteria enriched in patients with kidney rejection F) Differential abundance analysis of bacterial genera, with significant changes identified between non-rejection and rejection groups. Cliff’s Delta values indicate the direction and extent of these differences. Blue dots show genera increased in the non-rejection group and red increased in the rejection group.

Microbiome analysis indicated alterations prior to the rejection event. Pre-rejection microbiome composition was characterized by a distinct composition indicated by different clustering in the PCoA of Bray-Curtis dissimilarity (Fig 2C), and lower alpha-diversity, reaching significance for Simpson evenness (Fig 2D). Patients who rejected the transplanted kidney had lower level of known SCFA producers on genus level, like *Blautia*, *Clostridia*, or *Ruminococcus* torques group^38^. Interestingly, we observed an increase in bacteria typically found in CKD patients, like *Fusobacterium*^8^, and disease-associated genera^40^, such as *Strepcoccus*^39^ and *Porphyromonas*^41^ (Fig 2E).

Despite our efforts to closely match the rejection and non-rejection groups, a higher rate of viral infections was observed in the rejection group. We therefore carefully assessed the use of immunosuppressants and anti-infective medication (Supplemental Figures 5-7), pointing out two main differences, namely a lower number of patients on MPA at sampling date – likely due to the viral complications – and a lower number of patients with basiliximab for induction in the rejection group. Multivariate analysis revealed no difference in microbiome composition in rejection patients with or without basiliximab induction (Supplemental Figure 8), but MPA withdrawal showed a non-significant shift in microbiome composition. Therefore, we performed the genus level differential abundance analysis, including and excluding patients with MPA discontinuation, which closely correlated (R= 0.98, p< 0.001) (Supplemental Figure 9). This indicates that the observed differences were independent of basiliximab induction or MPA discontinuation, respectively.

### Lower SCFA production potential characterizes the pre-rejection microbiome

Metabolites produced by microbiota are known to influence and modulate host immune responses. To identify potential candidates, we analyzed functional capacities, predicted from our taxonomic data using PICRUSt2^32^, identifying functional pathways in the significantly altered KOs using GOmixer^33,42^. We observed an overall clustering of rejection and non-rejection patients in the PCA (Fig 3A). We found an enrichment in proteolytic fermentation, reactive nitrogen and oxygen species, and ammonia pathways in the rejection group (Fig 3B, C). Conversely, overall sugar and polysaccharide utilization and mucus degradation were enriched in microbiomes from non-rejection controls (Fig 3B, C). Matching to the reduced number of SCFA-producing genera we found a reduction in butyrate and acetate fermentation pathways (Fig 3B, C), again highlighting the reduction of SCFA production in stool samples preceding KT rejection (Fig 3B, C). Since SCFA and regulatory immune functions are closely linked, we aimed to confirm the reduction of SCFA production using qPCR measuring key enzymes for butyrate and propionate production. Overall, we found a significant reduction of butyryl-CoA:acetate CoA-transferase (*but*), a key enzyme for butyrate production, and methylmalonyl-CoA decarboxylase (*mmdA*), a key enzyme for propionate production (Fig 4A, B). Another central enzyme for butyrate synthesis, butyryl-CoA dehydrogenasee (*bcd*), was reduced without reaching significance (Fig 4A, B). Taken together, a key feature of the gut microbiome in samples from patients preceding KT rejection is a marked reduction of the potential to produce SCFA.

**Figure 3.**
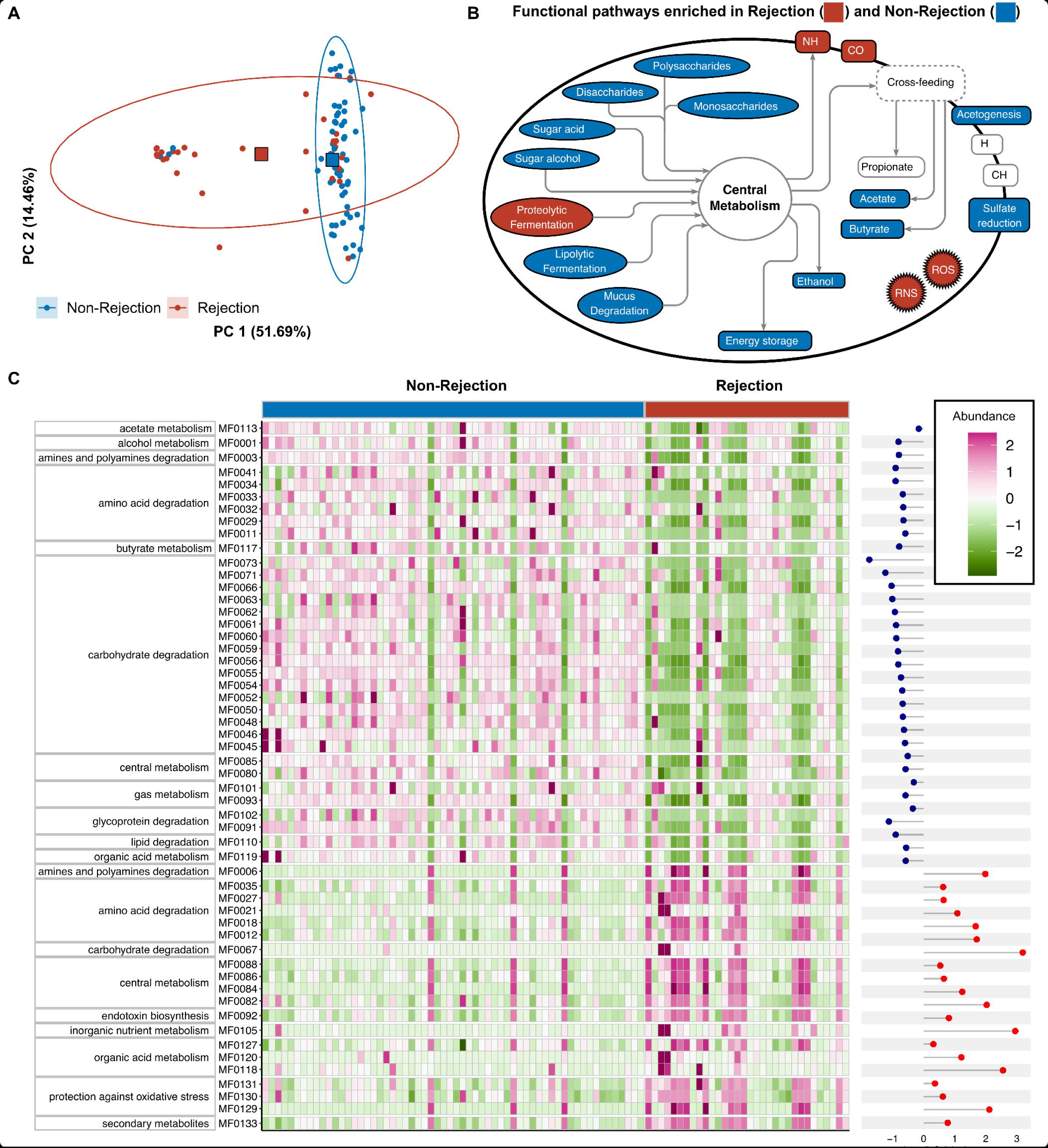
Fecal short-chain fatty acid production potential is reduced before kidney transplant rejection. Patients with available fecal samples prior to kidney rejection (Rejection group) for propensity score matched (1:2) to patients without kidney rejection (Non-Rejection group). Inference of functional data from taxonomic data using PICRUSt2 and mapping to pathways using GOmixer. A) Clustering of kidney transplant recipients based on the presence (red circles) or absence (blue circles) of graft rejection, illustrating differences in microbiome-derived functional potential. Squares represent centroids. B) Functional pathways highlight an increase in proteolytic fermentation and reactive nitrogen and oxygen species production pathways in patients who experienced graft rejection (red). Non-rejection samples show enhanced pathways related to sugar and polysaccharide metabolism, and mucus degradation (blue). C) Heatmap shows all 54 significant GOmixer modules, with pink indicating high predicted abundances and green representing low abundances across the 92 patients, divided into non-rejection and rejection. Blue dots denote negative log2 fold changes, indicating enhanced modules in non-rejection samples compared to rejection samples, while red dots represent positive log2 fold changes, signifying an increase in patients experiencing graft rejection.

**Figure 4.**
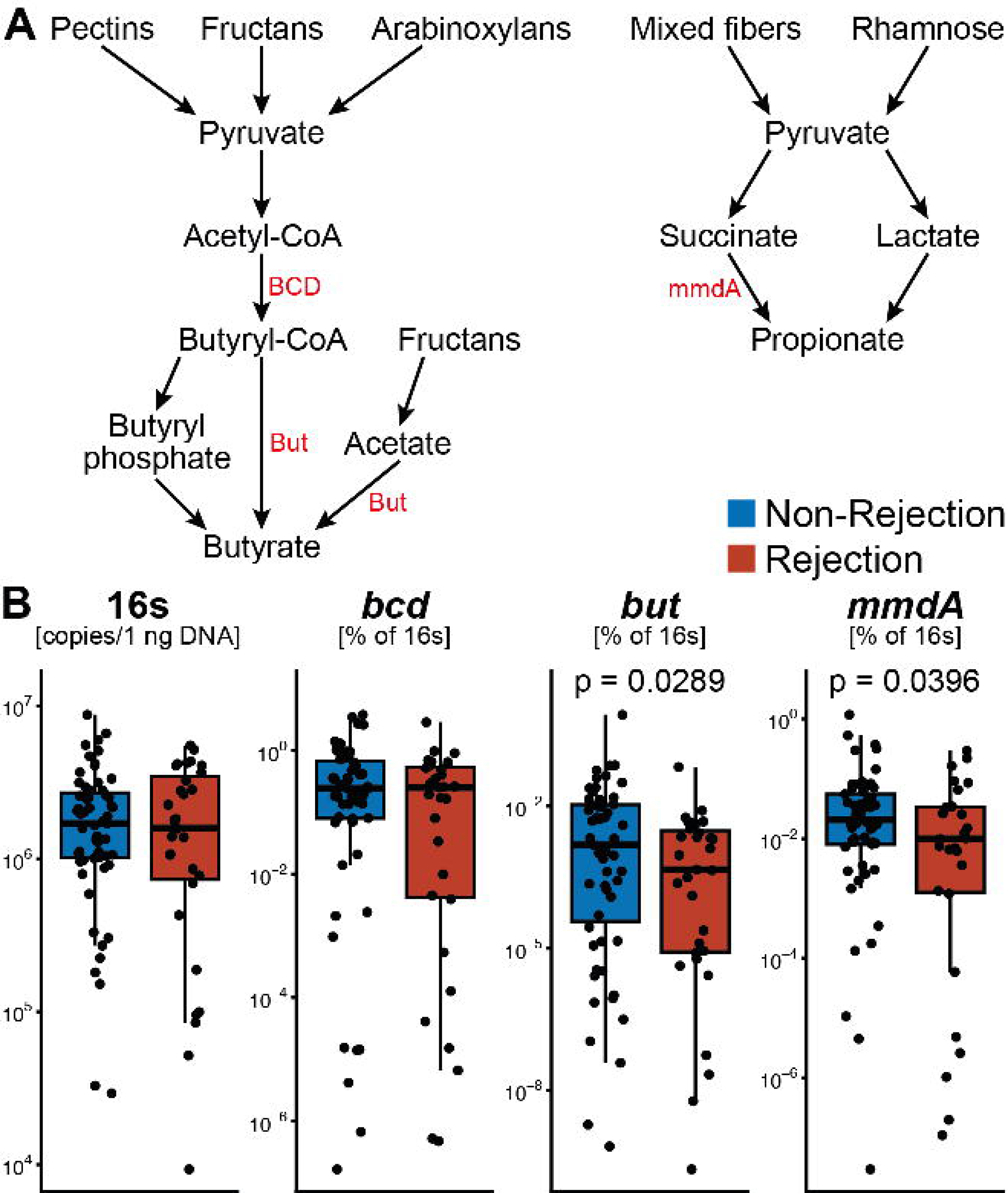
Decreased abundance of short-chain fatty acid producing enzymes in the microbiome before kidney transplant rejection. Patients with available fecal samples prior to kidney rejection (Rejection group) for propensity score matched (1:2) to patients without kidney rejection (Non-Rejection group). A) Bacterial nutrient metabolism leading to SCFA, specifically butyrate and propionate, production. Key bacterial enzymes highlighted in red. B) Abundance of bacterial enzymes for SCFA production were assessed by qPCR from fecal DNA and normalized to 16s content. While *bcd* was not significantly altered *but* and *mmdA* showed lower abundance in fecal samples from patients with subsequent kidney rejection compared to those without rejection.

### Microbiome alterations normalize post-rejection

As our initial analysis indicated a potential shift post-rejection, we analyzed longitudinal samples from our matched cohort within a timeframe of 90 to 1000 days after the first sample (n=21 rejection and n=54 non-rejection patients). PCoA using Bray-Curtis dissimilarity shows that the microbiome composition becomes more similar to the non-rejection control group in post-rejection samples (Fig 5A). Over a similar time frame the microbiome composition of non-rejection controls was stable. The same trend could be observed for the number of detected OTUs, Shannon diversity and Simpson evenness (Fig 5B). Next, we analyzed microbiome alterations post-rejection on genus level (Fig 5C). While the non-rejection control showed no significant alterations, most genera dysregulated preceding rejection were significantly changed in the opposite direction after the KT rejection event (Fig 5C). Especially known genera of SCFA production like *Blautia* and *Faecalibacterium* increased, while disease-associated genera like *Fusobacterium* and *Streptococcus* decreased (Fig 5D).

**Figure 5.**
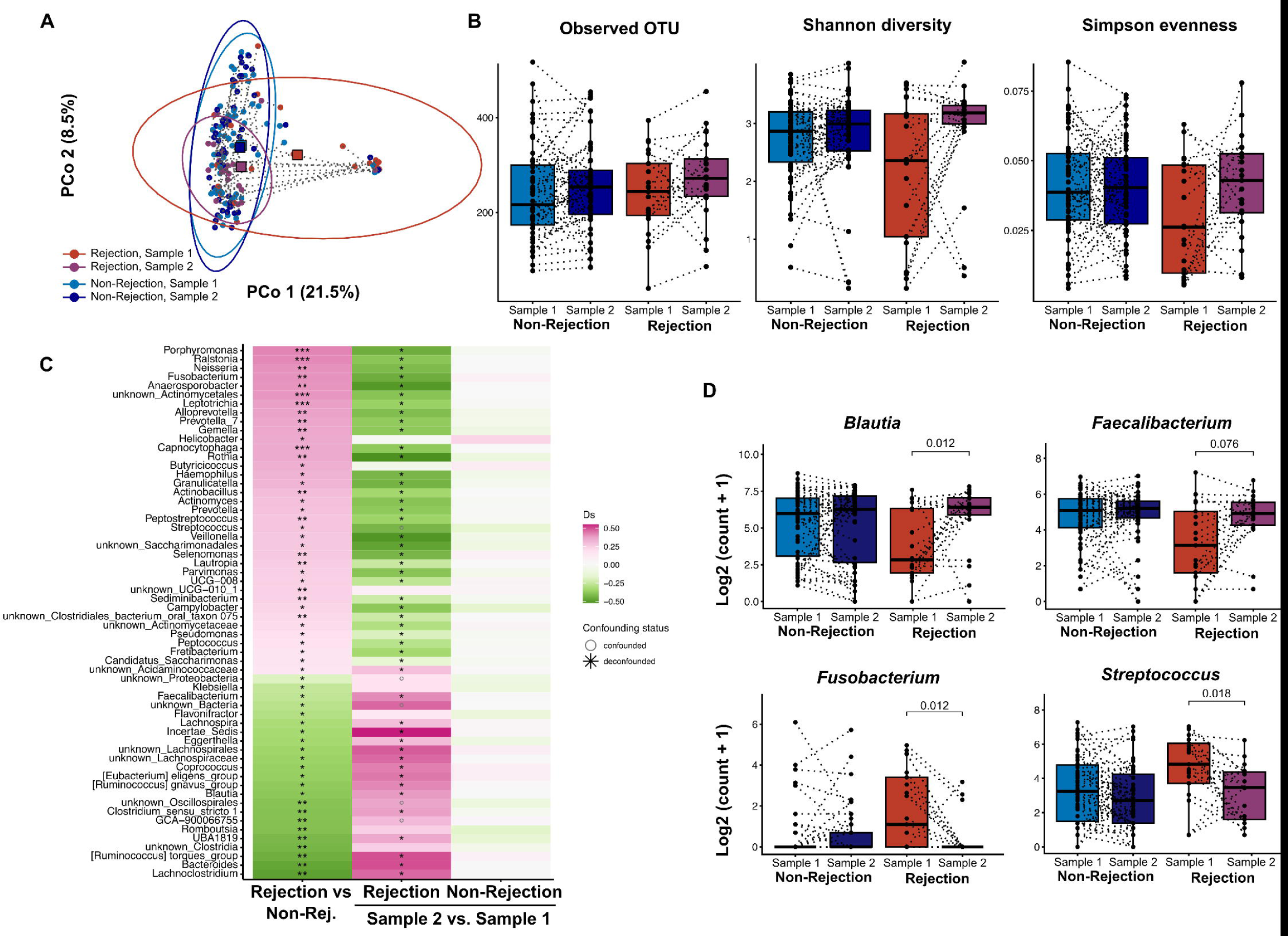
Microbiome composition normalizes post-kidney transplant rejection. Patients with available fecal samples (sample 1) prior to kidney rejection (Rejection group) and sample 2 90-1000 days post-rejection were analyzed. Propensity score matched patients without kidney rejection (Non-Rejection group) with two comparable samples were included as controls. A) Principal Coordinates Analysis (PCoA) of microbiome samples based on Bray-Curtis dissimilarity demonstrates a shift in the microbiome composition of rejection patients (red to purple) towards the non-rejection group (blue) over time, suggesting a post-rejection stabilization. Squares represent centroids. B) Microbial diversity measured by the number of detected OTU, Shannon diversity, and Simpson evenness. C) Heatmap comparison of genus-level alterations between pre- and post-rejection samples reveals significant directional changes in microbial populations after a rejection event, contrasting with the stable profiles observed in non-rejection controls. Pre-rejection comparison taken from Figure 3F. Asterisks indicate significance levels: FDR < 0.1 * FDR <0.01 ** FDR <0.001 ***. Confounded signals are shown as circles. Effect size is shown as Cliffs delta with pink enriched and green depleted. D) Boxplots show shifts at the genus level, of short-chain fatty acid producers, *Blautia* and *Faecalibacterium,* and disease-associated genera, *Fusobacterium* and *Streptococcus*.

Taken together, we observe a normalization of the pre-rejection microbiome towards the normal KT signature in a longitudinal follow-up analysis. One could speculate that a lack of normalization of the microbiome favors chronic rejections. Due to insufficient sample size, we could not further test this hypothesis in this study.

### The microbiome signature in kidney transplant rejection is a prolonged CKD signature

Lastly, we aimed to contextualize the microbiome alteration preceding KT rejection. We observed a partial overlap with a microbiome features of pediatric CKD patients recently published by us^8^. Therefore, we hypothesized that the microbiome alterations preceding KT rejection in parts reflects a prolonged CKD signature after KT. Therefore, we re-analyzed a recently published 16S rRNA gene sequencing dataset from CKD patients (n=217) and healthy controls (n=479)^26^. In line with our hypothesis, the KT rejection signature correlated with the CKD signature in the dataset from Ren et al. (for the 120 common genera, R= 0.19, p= 0.033) (Fig 6A). In particular, the overlap of both datasets also held true for the reduction of important SCFA producers like *Blautia* and *Faecalibacterium*, as well as for the increase in *Streptococcus* and *Fusobacterium*, with the latter not reaching significance in the CKD-HC comparison (Fig 6A, B). Lastly, we performed a targeted analysis of bacterial taxa captured by our *but*, *bcd* and *mmdA* assays. Overall, the abundance of butyrate and propionate producing taxa was lower in CKD patients mirroring the effect we observed in microbiome samples preceding KT rejection (Fig 6C).

**Figure 6.**
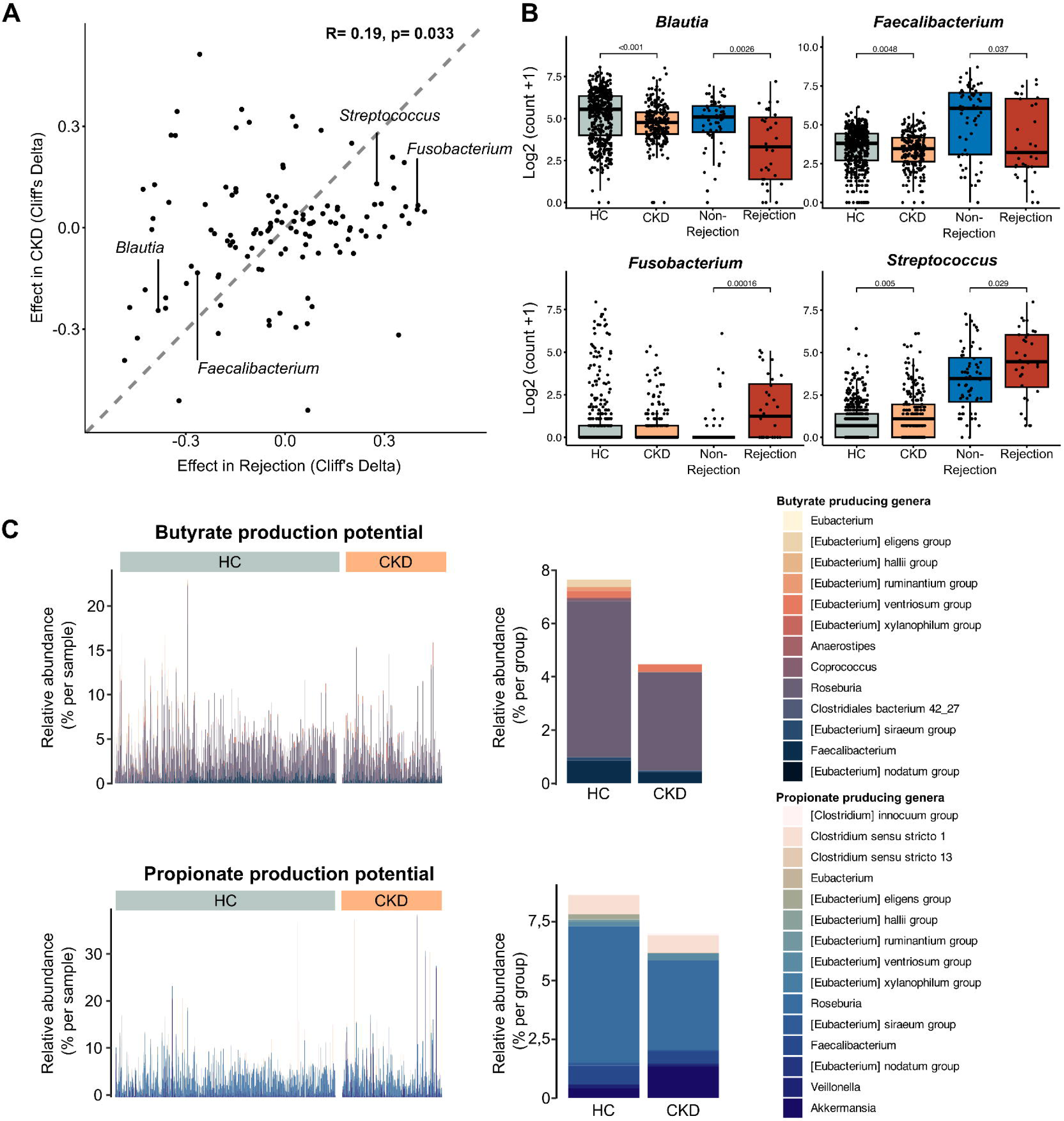
Microbiome composition in kidney rejection partly mirrors CKD-related microbiome alterations. Correlation between CKD microbial signatures and kidney transplant rejection preceding signatures. A) Scatter plot indicating the correlation between the microbial signatures observed before kidney transplant (KT) rejections and those seen in chronic kidney disease (CKD) patients, suggesting shared alterations. Dashed line indicates perfect overlap of effect sizes. B) Log transformed rarefied abundance of key SCFA-producing genera (*Blautia* and *Faecalibacterium*) and disease-associated genera (*Streptococcus* and *Fusobacterium*) across healthy controls (HC), CKD patients, and KT recipients preceding graft rejection or without graft rejection (propensity-score matched). C) Abundance of bacterial taxa involved in butyrate and propionate production between HC and CKD patients, reflecting a decrease in these beneficial taxa in CKD.

In aggregate, our data indicate that the pre-rejection signature we observed in our cohort might in part be a sustained CKD signature. Especially the lack of fiber fermenting, SCFA-producing bacteria is a key feature found in both disease states.

## 4. Discussion

In the present study, we investigate the gut microbiome in KT recipients and its relationship with allograft rejection. KT remains the best treatment option for patients with advanced and dialysis-dependent CKD, but graft availability is limited. Therefore, the improvement of graft survival and prevention of allograft rejection are paramount. For the first time, we describe compositional and functional differences in the microbiome in a representative cohort of 217 transplant patients with and without rejection, which are of potential prognostic and therapeutic value.

Our analysis of longitudinally collected fecal samples unveils a dynamic trajectory of microbiome recovery post KT, which undertakes a gradual shift towards a more stable and healthier microbiota composition^40^ over time. This gradual process is marked by the enrichment of bacterial taxa associated with short-chain fatty acid (SCFA) production (such as *Roseburia, Faecalibacterium*), and a loss of disease-associated taxa (such as *Streptococcus*). These regenerative microbiome shifts post KT are significantly perturbed in the case of graft rejection. Preceding the rejection event, we observed profound alterations in microbiome composition, characterized by a diminished diversity and underrepresentation of SCFA-producing bacterial populations. Notably, these alterations tend to normalize post KT rejection events. We consider these observations to be of potential functional relevance for allograft immunity due to the known immunomodulatory properties of SCFA reference^9,17^. Although our study was not designed to show causality, our results suggest that the microbiome is an important modulator of immunologic events after KT - an observation that may also have prognostic significance for the prevention of KT rejection and graft survival.

The observed gradual normalization of the microbiome composition after KT aligns for several bacterial taxa the normalization of known alterations in the gut microbiome of CKD patients. The gut microbiome of CKD patients is relatively well investigated. It is characterized by a lower diversity and a shift in metabolic output^6,8,26,43^. While the production of SCFA metabolites, recognized for their anti-inflammatory effects^9^, is reduced^8^, we and others have observed an increase in the production of microbiome-derived uremic toxins like TMA^44^, p-cresols^45^ and indoles^8,46^. Of note, the microbiome of CKD patients is characterized by an increase of bacterial taxa frequently linked to health-to-disease transition in large representative metagenomic studies^40,47^. In the present study, we observe a decrease of known pathogenic bacteria and increase in beneficial commensals over the course of more than three years post KT. Thus, the observed changes are reminiscent of a CKD-to-health transition in the microbiome. Dysbiotic microbiome states in KT patients have been described by others^10,13^. Swarte and colleagues demonstrated that lower gut microbial diversity in kidney and liver transplant recipients is associated with an increased overall mortality^13^. As graft survival is the most important predictor of overall medium- and long-term survival after KT^2^, we hypothesize that gut bacterial diversity may also be important for graft survival.

We demonstrate that alterations of the gut microbiome, including a reduced microbial diversity, and alterations in the abundance of more than 50 bacterial taxa, occur before graft rejection. Microbiome composition preceding graft rejection was characterized by several typical disease-associated species. Most prominent and similar to the analyzed CKD signature, we found an increase in *Streptococcus*. *Streptococcus spp.* were recently correlated to subclinical atherosclerotic lesions in a cohort of nearly 9000 patients^39^. Another genus found to be upregulated in rejection and CKD (although not reaching significance in the cohort we reanalyzed^26^ as compared to other studies^8^) was *Fusobacterium*. *Fusobacterium nucleatum* was recently described as a driver of uremic toxins production and CKD progression^43^, again underscoring the notion of a prolonged CKD signature in the microbiome as a potential risk factor for graft rejection. We and others have shown that SCFA are relevant bacterial metabolites for immune homeostasis in CKD as well as for other chronic diseases^8,17,48–50^. Using computational prediction of functional microbiome properties (PICRUSt2^32^), we show a significantly reduced potential of the gut microbiome to produce SCFA. PICRUSt2 predicts microbiome functions such as SCFA production based on the available genomic data and the inferred presence of genes from closely related taxa^32^. PICRUSt accuracy significantly decreases for microbes with fewer close relatives in reference databases^51^. Acknowledging this limitation, we validated the regulation of several key enzymes for SCFA synthesis using gene targeting qPCR assays. Therefore, the microbiome may be functionally implied in the development of graft rejection.

A recent meta-analysis concluded that immune cell therapies, including transfer of regulatory T cells (Treg), are a useful approach to reduce immunosuppression during KT^20^. The findings of our study and others suggest that SCFA warrant scientific attention as Treg-modulating bacterial metabolites. SCFA have been shown to influence the function and differentiation of Treg both through GPR signaling and HDAC inhibition^9,52^. Experimental transplantation models demonstrate the efficacy of SCFA treatments through induction of Treg. Kidney transplants showed a prolonged survival in animals fed a high fiber diet or directly the SCFA acetate^53^. Thus, the rapid attainment of a healthy SCFA production potential could be relevant for Treg function and the prevention of rejection events as well as other comorbidities found after KT^54^.

Two recent smaller studies similarly suggest alterations of the microbiome in patients with acute rejection (n=3 patients, including both TCMR and antibody-mediated rejection)^55^ or patients with antibody-mediated rejection (n=24)^56^. While the first study is severely limited by its low sample size, the second study – albeit focusing on ABMR compared to our focus on TCMR – described on a broad scale similar features like a reduced alpha diversity. Of note, the observed microbiome alterations in our study preceding rejection events normalized during longitudinal follow-up. Future studies are needed to investigate whether persistent microbiome alterations influence the risk for chronic rejection.

The circumstance that fecal samples were not consistently available from all patients at all timepoints in our multi-center cohort limits the validity of our study. However, in non-rejection patients a minimum sample number of two stool samples per patient was not undercut, which enabled the longitudinal character of our study. Despite this, our findings clearly indicate a gradual shift towards a more stable microbiome state post-transplantation. A further limitation is the lack of metabolomic measurements in our study, which we replaced with specific quantifications of the microbial enzyme composition. Finally, the study design does not allow causal statements on the mechanistic significance of microbiome changes for rejection. However, our conclusions regarding the role of SCFA for Treg function are broadly based and supported by published experimental data. Future studies should consider functional data on immune cells and their relationship to microbiome and rejection to validate our conclusions.

Taken together, we demonstrate a disrupted microbiome recovery post KT as a novel modifying factor in graft survival. This is to our knowledge the first study indicating that microbiome alterations and perturbation of microbial metabolism precede graft rejection. More studies are needed to decipher the interaction of SCFA and Treg in KT patients and to test the potential of microbiome-targeting interventions before and after KT to improve long term graft survival.

## Funding

This study was conducted with resources provided by the DZIF transplant cohort e.V. (https://www.dzif.de/en/working-group/transplant-cohort), support code TTU 07.701, and additional DZIF funding under TTU 07.916 to M. Gerhard, D. Schinderl and J. Holle. J Holle was supported by the Else Kröner-Fresenius Stiftung (2023_EKEA.127). N. Wilck was supported by the European Research Council under the European Union’s Horizon 2020 research and innovation program grant 852796 and by the Corona Foundation in the German Stifterverband. H. Bartolomaeus and N. Wilck were supported by the BMBF, Foerderkennzeichen 01EJ2202A (TAhRget consortium).

## Data Availability

Deidentified metagenomic sequencing data for samples of the Transplant Cohort of the DZIF can be accessed from the European Nucleotide Archive under accession number PRJNA1106540 (https://www.ebi.ac.uk/ena/browser/view/PRJNA1106540). Access to pseudonymized phenotype data requires approval from the scientific steering committee of the DZIF Transplant cohort. The source code and the summary data underlying all figures used to generate the results for the analysis are available at https://github.com/rosareitmeir/DZIF-Tx-Cohort-Data-Cleaning-and-Statistical-Analysis.

## Acknowledgement

We thank the members of the German Center of Infection Research Transplant Cohort, in particular the members of the executive board and the scientific steering committee: Dr. Susanne Delecluse, Dr. Tina Ganzenmüller, Dr. Bärbel Fösel, Dr, Caroline Klett-Tammen, Dr. Thomas F. Schulz, Dr. Uwe Heemann, Dr. Thomas Giese, Dr. Philip Ehlermann, Dr. Rasmus Rivinius, Dr. Uta Merle, Dr. Peter Lang, Dr. Martin Hildebrandt, Dr. Christine S. Falk, Dr. Burkhard Tönshoff, Dr. Anette Melk, Dr. Thomas Illig, Dr. Berit Lange, and Dr. Thomas Iftner.

## Supplemental Material

**Supplemental Figure 1. Study design.** Flow chart of the study design with inclusion and exclusion of patients and final sample number.

**Supplemental Figure 2. Observed OTU in rejection (red) and non-rejection (blue) patients.** Horizontal lines indicate the number of detected OTU, shaded area is the 95% confidence interval. Vertical lines on top of the x-axis indicate rejection events.

**Supplemental Figure 3. Impact of rejection state on gut microbiome composition.** Analysis of fecal samples from patients experiencing kidney rejection at any time compared to patients never experiencing graft rejection. Heatmap showing associations of bacterial genera with kidney transplant rejection. Meta-variables with significant association are shown. Stars indicate de-confounded significant associations; grey dots indicate confounded associations. °/*FDR< 0.1, °/**FDR< 0.01, °°/***FDR< 0.001.

**Supplemental Figure 4: Matching of rejection and non-rejection patients.** Plots indicate metavariables before (left) and after propensity score matching (right). Grey bars/ histogram indicate non-rejection before (left) and after (right) propensity score matching. Rejection is shown in black.

**Supplemental Figure 5: Immunosuppressive treatment at the time of transplantation (A) and at the time of fecal sampling (B).** Fecal samples were analysed from patients before experiencing graft rejection (Rejection, red) and propensity score matched (1:2) controls (normal progress, blue). Bar plots indicate medication at A) transplantation and B) fecal sampling.

**Supplemental Figure 6. Tacrolimus plasma levels in matched rejection and non-rejection patients.** A) Tacrolimus plasma levels of n=92 patients (matched cohort) for the time between transplantation and sample collection before rejection event (n=121 data points of 51 patients for non-rejection and 51 data points of 23 patients for rejection group). B) Difference of target to actual values of Tacrolimus. Target values have been determined by the respective clinical recommendations after kidney transplantation.

**Supplemental Figure 7: Anti-infective medications used between kidney transplantation and fecal sampling.** Anti-infective medication used between kidney transplantation and fecal sampling for the matched cohort (normal progress, blue; rejection, red). Medication used both preventive and therapeutic is shown.

**Supplemental Figure 8: Impact of basiliximab induction therapy on the gut microbiome composition.** PCoA on Bray-Curtis Dissimilarity of the 16S amplicon sequencing in rejection patients with (brown) and without (orange) basiliximab use during induction. Statistical testing using PERMANOVA shows no difference.

**Supplemental Figure 9: Impact of mycophenole (MPA) withdrawal on the gut microbiome composition.** A) PCoA on Bray-Curtis Dissimilarity of the 16S amplicon sequencing in rejection patients with (light green) and without (dark green) MPA at fecal sampling. Statistical testing using PERMANOVA shows no difference, although some distinct clustering can be seen. B) Correlation of effect sizes in the whole rejection cohort (x-axis) and the cohort excluding patients with MPA withdrawing (y-axis). Axis shows Cliff’s deltas for all detected genera in the gut microbiome. Spearman correlations, individual significances for genera using univariate testing in metadeconfoundR.

## Supplemental Tables

**Supplemental Table 1:** Clinical variables used for deconfounding of microbiome features

**Supplemental Table 2:** Primers used for 16S sequencing.

**Supplemental Table 3:** Bacterial species included to the gene targeting assays for butyrate and propionate synthesis and corresponding primer sequences.

## Notes

### Competing Interest Statement

The authors have declared no competing interest.

### Author Declarations

Ethics approval was obtained from all participating centers (University of Heidelberg, Heidelberg, Germany: #S-585/2013; Technical University of Munich (TUM), TUM School of Medicine and Health, Munich, Germany: #5926/13; Klinikum der Universitat Munchen, Munich, Germany: #380-15; University Hospital Tubingen, Tubingen, Germany: #327/2014BO1), and all participants provided written informed consent. The experimental analysis of fecal samples, including metadata analysis, was approved by the ethics board of Charite - Universitatsmedizin Berlin, Berlin, Germany (EA2/208/21).

## References

1. Webster AC, Nagler EV, Morton RL, Masson P. Chronic Kidney Disease. Lancet. Mar 25 2017;389(10075):1238-1252. doi:10.1016/S0140-6736(16)32064-5

2. Hariharan S, Israni AK, Danovitch G. Long-Term Survival after Kidney Transplantation. N Engl J Med. Aug 19 2021;385(8):729–743. doi:10.1056/NEJMra2014530

3. Cooper JE. Evaluation and Treatment of Acute Rejection in Kidney Allografts. Clin J Am Soc Nephrol. Mar 6 2020;15(3):430–438. doi:10.2215/CJN.11991019

4. Belkaid Y, Hand TW. Role of the microbiota in immunity and inflammation. Cell. Mar 27 2014;157(1):121–141. doi:10.1016/j.cell.2014.03.011

5. Belkaid Y, Harrison OJ. Homeostatic Immunity and the Microbiota. Immunity. Apr 18 2017;46(4):562-576. doi:10.1016/j.immuni.2017.04.008

6. Behrens F, Bartolomaeus H, Wilck N, Holle J. Gut-immune axis and cardiovascular risk in chronic kidney disease. Clin Kidney J. Jan 2024;17(1):sfad303. doi:10.1093/ckj/sfad303

7. Schlender J, Behrens F, McParland V, et al. Bacterial metabolites and cardiovascular risk in children with chronic kidney disease. Mol Cell Pediatr. Oct 22 2021;8(1):17. doi:10.1186/s40348-021-00126-8

8. Holle J, Bartolomaeus H, Lober U, et al. Inflammation in Children with CKD Linked to Gut Dysbiosis and Metabolite Imbalance. J Am Soc Nephrol. Dec 2022;33(12):2259–2275. doi:10.1681/ASN.2022030378

9. Smith PM, Howitt MR, Panikov N, et al. The microbial metabolites, short-chain fatty acids, regulate colonic Treg cell homeostasis. Science. Aug 2 2013;341(6145):569-573. doi:10.1126/science.1241165

10. Swarte JC, Douwes RM, Hu S, et al. Characteristics and Dysbiosis of the Gut Microbiome in Renal Transplant Recipients. J Clin Med. Feb 1 2020;9(2)doi:10.3390/jcm9020386

11. Lee JR, Magruder M, Zhang L, et al. Gut microbiota dysbiosis and diarrhea in kidney transplant recipients. Am J Transplant. Feb 2019;19(2):488–500. doi:10.1111/ajt.14974

12. Fricke WF, Maddox C, Song Y, Bromberg JS. Human microbiota characterization in the course of renal transplantation. Am J Transplant. Feb 2014;14(2):416–427. doi:10.1111/ajt.12588

13. Swarte JC, Li Y, Hu S, et al. Gut microbiome dysbiosis is associated with increased mortality after solid organ transplantation. Sci Transl Med. Aug 31 2022;14(660):eabn7566. doi:10.1126/scitranslmed.abn7566

14. Lei YM, Chen L, Wang Y, et al. The composition of the microbiota modulates allograft rejection. J Clin Invest. Jul 1 2016;126(7):2736–2744. doi:10.1172/JCI85295

15. Sato Y, Yanagita M. Immunology of the ageing kidney. Nature reviews Nephrology. Oct 2019;15(10):625–640. doi:10.1038/s41581-019-0185-9

16. Wilck N, Matus MG, Kearney SM, et al. Salt-responsive gut commensal modulates TH17 axis and disease. Nature. Nov 30 2017;551(7682):585-589. doi:10.1038/nature24628

17. Bartolomaeus H, Balogh A, Yakoub M, et al. Short-Chain Fatty Acid Propionate Protects From Hypertensive Cardiovascular Damage. Circulation. Mar 12 2019;139(11):1407–1421. doi:10.1161/CIRCULATIONAHA.118.036652

18. Wu H, Singer J, Kwan TK, et al. Gut Microbial Metabolites Induce Donor-Specific Tolerance of Kidney Allografts through Induction of T Regulatory Cells by Short-Chain Fatty Acids. J Am Soc Nephrol. Jun 1 2020;doi:10.1681/ASN.2019080852

19. Singer J, Li YJ, Ying T, et al. Protocol for a pilot single-centre, parallel-arm, randomised controlled trial of dietary inulin to improve gut health in solid organ transplantation: the DIGEST study. BMJ Open. 2021;11(4)

20. Sawitzki B, Harden PN, Reinke P, et al. Regulatory cell therapy in kidney transplantation (The ONE Study): a harmonised design and analysis of seven non-randomised, single-arm, phase 1/2A trials. Lancet. May 23 2020;395(10237):1627–1639. doi:10.1016/S0140-6736(20)30167-7

21. Karch A, Schindler D, Kuhn-Steven A, et al. The transplant cohort of the German center for infection research (DZIF Tx-Cohort): study design and baseline characteristics. Eur J Epidemiol. Feb 2021;36(2):233–241. doi:10.1007/s10654-020-00715-3

22. Deutsche Stiftung Organtransplantation (German Organ Procurement Organization). Nierentransplantation: Grafiken zum Tätigkeitsbericht 2022 nach § 11 Abs. 5 TPG. 2022.

23. Kasiske BL, Zeier MG, Chapman JR, et al. KDIGO clinical practice guideline for the care of kidney transplant recipients: a summary. Kidney Int. Feb 2010;77:299-311. doi:10.1038/ki.2009.377

24. Ho DE, Imai K, King G, Stuart EA. Matching as Nonparametric Preprocessing for Reducing Model Dependence in Parametric Causal Inference. Political Analysis. 2007;15(3):199–236. doi:10.1093/pan/mpl013

25. Loupy A, Haas M, Roufosse C, et al. The Banff 2019 Kidney Meeting Report (I): Updates on and clarification of criteria for T cell-and antibody-mediated rejection. Am J Transplant. Sep 2020;20(9):2318-2331. doi:10.1111/ajt.15898

26. Ren Z, Fan Y, Li A, et al. Alterations of the Human Gut Microbiome in Chronic Kidney Disease. Adv Sci (Weinh). Oct 2020;7(20):2001936. doi:10.1002/advs.202001936

27. Lahti L, Sudarshan S. microbiome R package. http://microbiome.github.io

28. McMurdie PJ, Holmes S. phyloseq: An R Package for Reproducible Interactive Analysis and Graphics of Microbiome Census Data. PLOS ONE. 2013;8(4):e61217. doi:10.1371/journal.pone.0061217

29. Oksanen J, Blanchet FG, Kindt R, et al. Community ecology package. R package version. 2013;2(0):321–326.

30. Forslund SK, Chakaroun R, Zimmermann-Kogadeeva M, et al. Combinatorial, additive and dose-dependent drug-microbiome associations. Nature. Dec 2021;600(7889):500-505. doi:10.1038/s41586-021-04177-9

31. Chen C-Y, Lӧber U, Forslund SK. LongDat: an R package for covariate-sensitive longitudinal analysis of high-dimensional data. Bioinformatics Advances. 2023;doi:10.1093/bioadv/vbad063

32. Douglas GM, Maffei VJ, Zaneveld JR, et al. PICRUSt2 for prediction of metagenome functions. Nature Biotechnology. 2020/06/01 2020;38(6):685-688. doi:10.1038/s41587-020-0548-6

33. Darzi Y, Falony G, Vieira-Silva S, Raes J. Towards biome-specific analysis of meta-omics data. ISME J. May 2016;10(5):1025–1028. doi:10.1038/ismej.2015.188

34. Notting F, Pirovano W, Sybesma W, Kort R. The butyrate-producing and spore-forming bacterial genus Coprococcus as a potential biomarker for neurological disorders. Gut Microbiome. 2023;4:e16. e16. doi:10.1017/gmb.2023.14

35. Zhang J, Song L, Wang Y, et al. Beneficial effect of butyrate-producing Lachnospiraceae on stress-induced visceral hypersensitivity in rats. J Gastroenterol Hepatol. Aug 2019;34(8):1368–1376. doi:10.1111/jgh.14536

36. Kasahara K, Krautkramer KA, Org E, et al. Interactions between Roseburia intestinalis and diet modulate atherogenesis in a murine model. Nat Microbiol. Dec 2018;3(12):1461–1471. doi:10.1038/s41564-018-0272-x

37. Lenoir M, Martín R, Torres-Maravilla E, et al. Butyrate mediates anti-inflammatory effects of Faecalibacterium prausnitzii in intestinal epithelial cells through Dact3. Gut Microbes. Nov 9 2020;12(1):1–16. doi:10.1080/19490976.2020.1826748

38. Reichardt N, Duncan SH, Young P, et al. Phylogenetic distribution of three pathways for propionate production within the human gut microbiota. ISME J. Jun 2014;8(6):1323–1335. doi:10.1038/ismej.2014.14

39. Sayols-Baixeras S, Dekkers KF, Baldanzi G, et al. Streptococcus Species Abundance in the Gut Is Linked to Subclinical Coronary Atherosclerosis in 8973 Participants From the SCAPIS Cohort. Circulation. 2023;148(6):459–472. doi:doi:10.1161/CIRCULATIONAHA.123.063914

40. Gupta VK, Kim M, Bakshi U, et al. A predictive index for health status using species-level gut microbiome profiling. Nat Commun. Sep 15 2020;11(1):4635. doi:10.1038/s41467-020-18476-8

41. Stanisic D, Jeremic N, Singh M, Pushpakumar S, Mokshagundam SPL, Tyagi SC. Porphyromonas gingivalis induces cardiovascular dysfunction. Can J Physiol Pharmacol. Aug 1 2023;101(8):413–424. doi:10.1139/cjpp-2022-0392

42. Vieira-Silva S, Falony G, Darzi Y, et al. Species-function relationships shape ecological properties of the human gut microbiome. Nat Microbiol. Jun 13 2016;1(8):16088. doi:10.1038/nmicrobiol.2016.88

43. Wang X, Yang S, Li S, et al. Aberrant gut microbiota alters host metabolome and impacts renal failure in humans and rodents. Gut. Dec 2020;69(12):2131–2142. doi:10.1136/gutjnl-2019-319766

44. Stubbs JR, House JA, Ocque AJ, et al. Serum Trimethylamine-N-Oxide is Elevated in CKD and Correlates with Coronary Atherosclerosis Burden. J Am Soc Nephrol. Jan 2016;27(1):305–313. doi:10.1681/ASN.2014111063

45. Opdebeeck B, Maudsley S, Azmi A, et al. Indoxyl Sulfate and p-Cresyl Sulfate Promote Vascular Calcification and Associate with Glucose Intolerance. J Am Soc Nephrol. May 2019;30(5):751–766. doi:10.1681/ASN.2018060609

46. Holle J, Kirchner M, Okun J, et al. Serum indoxyl sulfate concentrations associate with progression of chronic kidney disease in children. PLoS One. 2020;15(10):e0240446. doi:10.1371/journal.pone.0240446

47. Gacesa R, Kurilshikov A, Vich Vila A, et al. Environmental factors shaping the gut microbiome in a Dutch population. Nature. 2022/04/01 2022;604(7907):732-739. doi:10.1038/s41586-022-04567-7

48. Avery EG, Bartolomaeus H, Rauch A, et al. Quantifying the impact of gut microbiota on inflammation and hypertensive organ damage. Cardiovasc Res. Jul 29 2022;doi:10.1093/cvr/cvac121

49. Sanna S, van Zuydam NR, Mahajan A, et al. Causal relationships among the gut microbiome, short-chain fatty acids and metabolic diseases. Nat Genet. Apr 2019;51(4):600–605. doi:10.1038/s41588-019-0350-x

50. Kim CH. Complex regulatory effects of gut microbial short-chain fatty acids on immune tolerance and autoimmunity. Cellular & Molecular Immunology. 2023/04/01 2023;20(4):341-350. doi:10.1038/s41423-023-00987-1

51. Langille MGI, Zaneveld J, Caporaso JG, et al. Predictive functional profiling of microbial communities using 16S rRNA marker gene sequences. Nature Biotechnology. 2013/09/01 2013;31(9):814-821. doi:10.1038/nbt.2676

52. Arpaia N, Campbell C, Fan X, et al. Metabolites produced by commensal bacteria promote peripheral regulatory T-cell generation. Nature. Dec 19 2013;504(7480):451-455. doi:10.1038/nature12726

53. Wu H, Singer J, Kwan TK, et al. Gut Microbial Metabolites Induce Donor-Specific Tolerance of Kidney Allografts through Induction of T Regulatory Cells by Short-Chain Fatty Acids. Journal of the American Society of Nephrology. 2020;31(7)

54. Rangaswami J, Mathew RO, Parasuraman R, et al. Cardiovascular disease in the kidney transplant recipient: epidemiology, diagnosis and management strategies. Nephrology Dialysis Transplantation. 2019;34(5):760–773. doi:10.1093/ndt/gfz053

55. Lee JR, Muthukumar T, Dadhania D, et al. Gut microbial community structure and complications after kidney transplantation: a pilot study. Transplantation. Oct 15 2014;98(7):697-705. doi:10.1097/tp.0000000000000370

56. Wang J, Li X, Wu X, et al. Gut microbiota alterations associated with antibody-mediated rejection after kidney transplantation. Appl Microbiol Biotechnol. Mar 2021;105(6):2473–2484. doi:10.1007/s00253-020-11069-x

