## Supplemental Material for "Gut microbiome alterations precede graft rejection in kidney transplantation patients"

Holle J*, Reitmeir R*, et al.

**Supplemental Methods**

1. **Clinical Data Preparation**

The clinical data preparation was done using R (4.03).

Viral Infections: All entries of viral infections and complications were counted as one, except infections labeled as “suspected infection” or “detection of antibodies without signs of acute disease”. Infections are considered as separate occurrences if the distance between diagnoses is at least five days.

Bacterial Infections and Antibiotic Treatment: Bacterial Infections and complications labeled as suspected, probable, or possible disease are excluded. Only infections with a confirmed diagnosis or indicated by antibiotic treatments are counted. A minimum five-day interval is required between distinct infections. The administered antibiotics are binned into 15 broader antibiotic classes and further summarized in total number of taken antibiotics and number of distinct antibiotic classes.

The processing of all clinical variables, including the immunosuppressive drug regimen, viral and bacterial infections, antibiotic and prophylaxis treatments, blood and urine values, and further baseline characteristics, is accessible via GitHub: https://github.com/rosareitmeir/DZIF-Tx-Cohort-Data-Cleaning-and-Statistical-Analysis.

2. **Collection of fecal samples and DNA**

The samples were collected following established guidelines as described previously^1^. Stool samples were received in stool collection tubes containing DNA stabilizer (from Invitek Molecular) and transported on dry ice. For DNA isolation, 1200 µl of stool sample combined with DNA Stabilizer buffer and 5 Zirconia Beads II from the PSP Spin Stool DNA Basic Kit (Stratec, Invitek Molecular) were transferred to a 2 ml microcentrifuge tube. The DNA isolation was performed following the manufacturers. To enrich the bacterial DNA, we subjected the samples to an additional incubation step at 95 °C for 10 minutes, shaking at 900 rpm in a thermomixer, and then the protocol was followed as per the instructions. Finally, DNA was eluted into a 1.5 ml microcentrifuge tube with 200 µl of elution Buffer (preheated to 70 °C). The DNA concentration was measured using a NanoDrop, and the samples were stored at -80 °C until the next steps.

3. **16S Amplicon sequencing**

To target bacterial DNA, the V3/V4 region of the 16S rRNA gene was amplified in 25 cycles from 2µl of aliquoted working stocks of DNA using primers 341F-ovh and 785R-ovh (Supplemental Table 1)^2^. AMPure XP magnetic beads (Beckmann Coulter Life Sciences, Germany) were used for the purification of the amplicons according to Illumina’s 16S Metagenomic Sequencing Library Preparation guide. Samples were indexed with Nextera XT indices (Illumina) in a paired fashion for 8 cycles of PCR, followed by a second purification with AMPure XP beads. Indexed samples were pooled in an equimolar amount (4nM), adjusted to a final concentration of 4 pM, and sequenced on a MiSeq system (Illumina) in a paired-end reaction of 600 cycles using the MiSeq reagent kit v3 (Illumina). A 20% (v/v) spike-in of the PhiX standard library at 4 pM was additionally included. As a control to check for artifacts, a single negative control (PCR with nuclease-free water as template), as well as a positive control using a mock community (ZymoBIOMICS, No. D6300, Freiburg, Germany), were included throughout each sequencing run.

Sequencing reads were cleaned for host contamination using BBMap, aligning to human reference genome (HG38 GRCh38) masked for regions homologous to bacterial genes using the Progenomes2 database to filter out human reads. Parameters were adjusted for high specificity (95% identity).

Operational Taxonomic Units (OTUs) were defined using LotuS 2.16, with UPARSE algorithm for de novo sequence clustering. Sequences were quality controlled and clustered, specifically tailored for MiSeq data. The Silva 16S rRNA gene SSU database (version 138.1) was employed for taxonomic assignments. The analysis also included checks for chimeras using RDP classifier.

4. **Gene targeting assay (quantitative PCR)**

To measure the potential of the gut microbiome to produce the SCFA butyrate and propionate, gene targeting qPCR assays (GTA) to quantify abundance of key enzymes for butyrate, propionate and acetate to propionate synthesis were developed. The genes encoding butyryl-CoA dehydrogenasee (*bcd*), butyryl-CoA:acetate CoA-transferase (*but*) and methylmalonyl-CoA decarboxylase (*mmdA*) in various main SCFA producing bacterial strains were targeted with minimally degenerate primers.

Standard curve qPCR was performed on a Quantstudio 5 real-time PCR system (Thermofisher) using *Power*Up SYBR green qPCR master mix (Thermofisher) and primers shown in Supplemental Table 2 and 3. In brief, reactions were run at 50°C for 2 min, 95°C for 2 min, followed by 40 cycles of 95°C for 2 s and 60°C for 30 s, with 6.66ng DNA and 250nM primers in 5µl reaction volume (measured in triplicates). Standards ranging from 10^8^-10^2^ copies/µl were generated using microbial community DNA mix (Sigma-Aldrich), Microbial DNA standard from Escherichia coli (Sigma-Aldrich) or previously isolated fecal bacterial DNA and run in parallel to determine the copies of each target gene within the samples. Standard curve 16s rRNA gene qPCR was performed as a quantification of bacterial load and target genes were normalized to the copies of 16s rRNA within each sample.

5. **Re-analysis of the CKD dataset**

16S rRNA gene amplicon sequencing data from an existing CKD cohort^3^, obtained from 696 samples (217 CKD patients and 469 healthy controls), were accessed from the Sequence Read Archive database (bioproject accession number PRJNA562327). The data was then analyzed using the LotuS2 pipeline^424^. The pipeline includes sequence quality filtering, read merging, adapter and primer removal, chimera removal, clustering, and taxonomic classification based on the SILVA (v138) database^5^. After quality control, we performed (i) targeted analysis of the abundance of genera found in the matched rejection-normal progress cohort and (ii) the abundance of typical butyrate and propionate producing bacteria was analyzed, focusing on the same taxa utilized in the design of the GTA (Supplemental Table 2).

**Supplemental Figures**

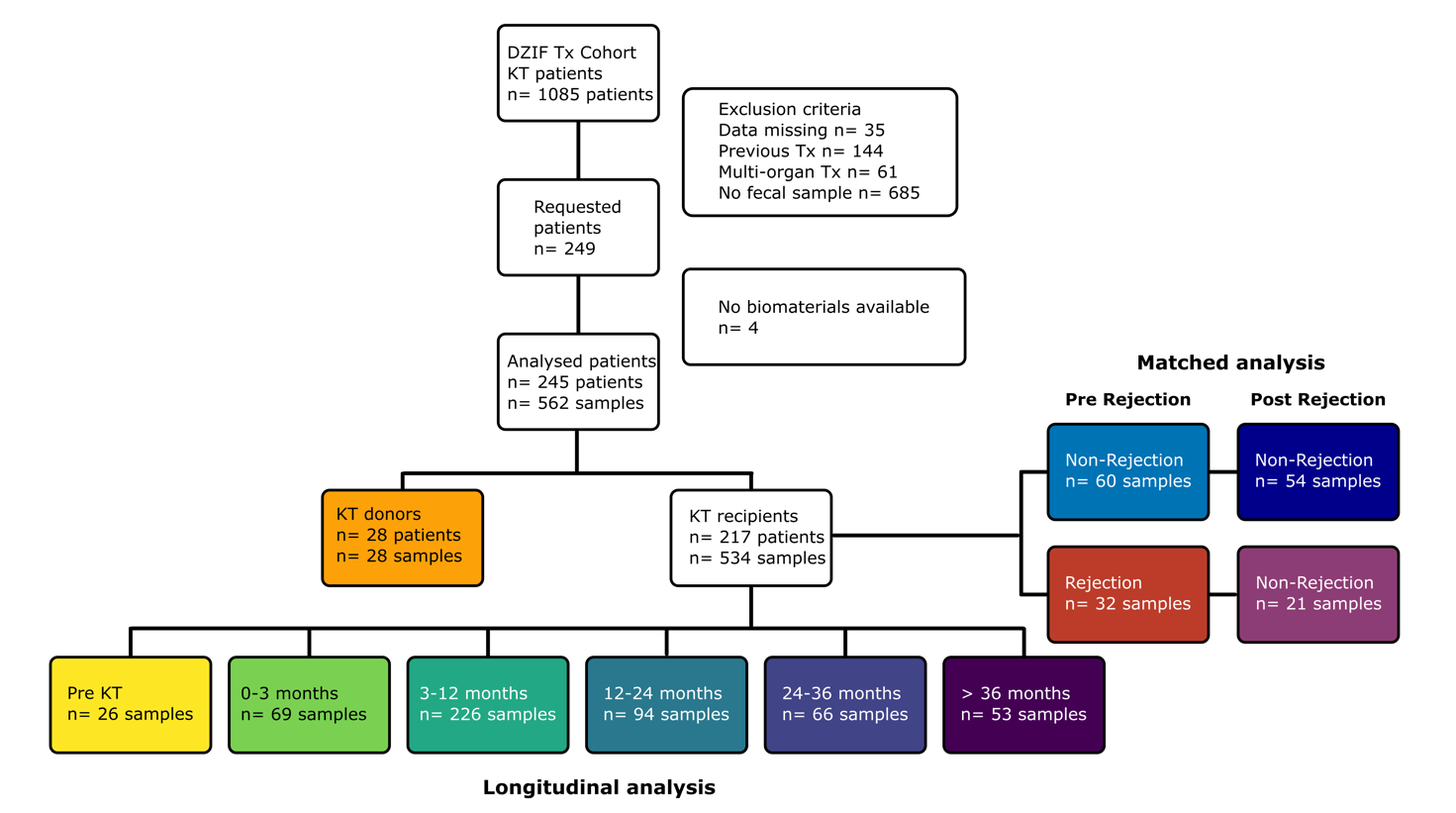

**Supplemental Figure 1. Study design.** Flow chart of the study design with inclusion and exclusion of patients and final sample number.

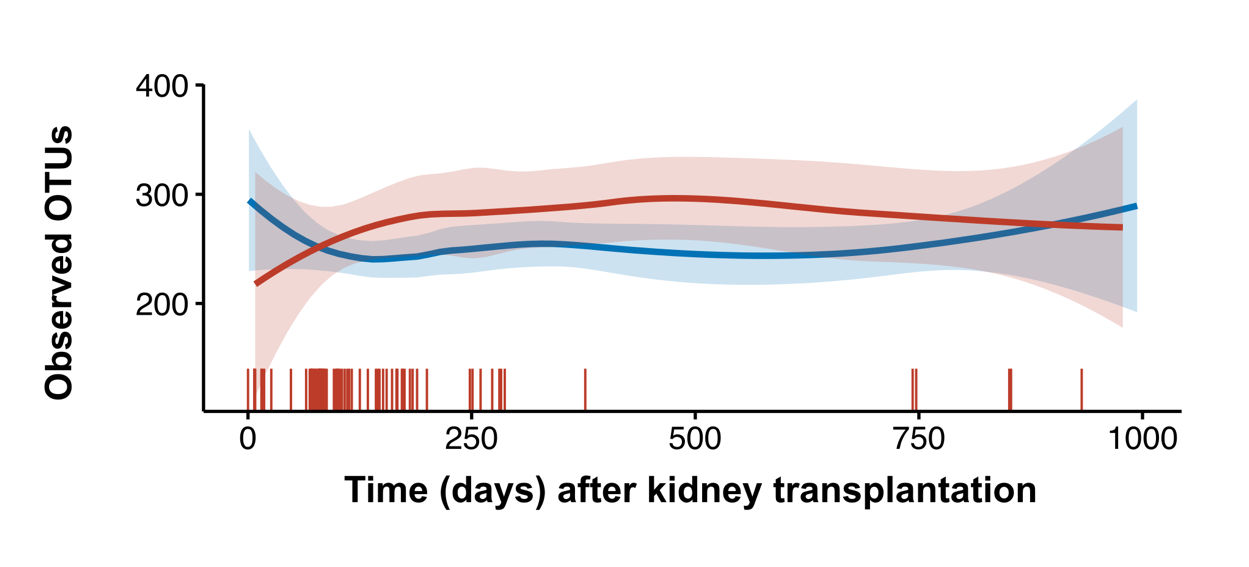

**Supplemental Figure 2. Observed OTU in rejection (red) and non-rejection (blue) patients.** Horizontal lines indicate the number of detected OTU, shaded area is the 95% confidence interval. Vertical lines on top of the x-axis indicate rejection events.

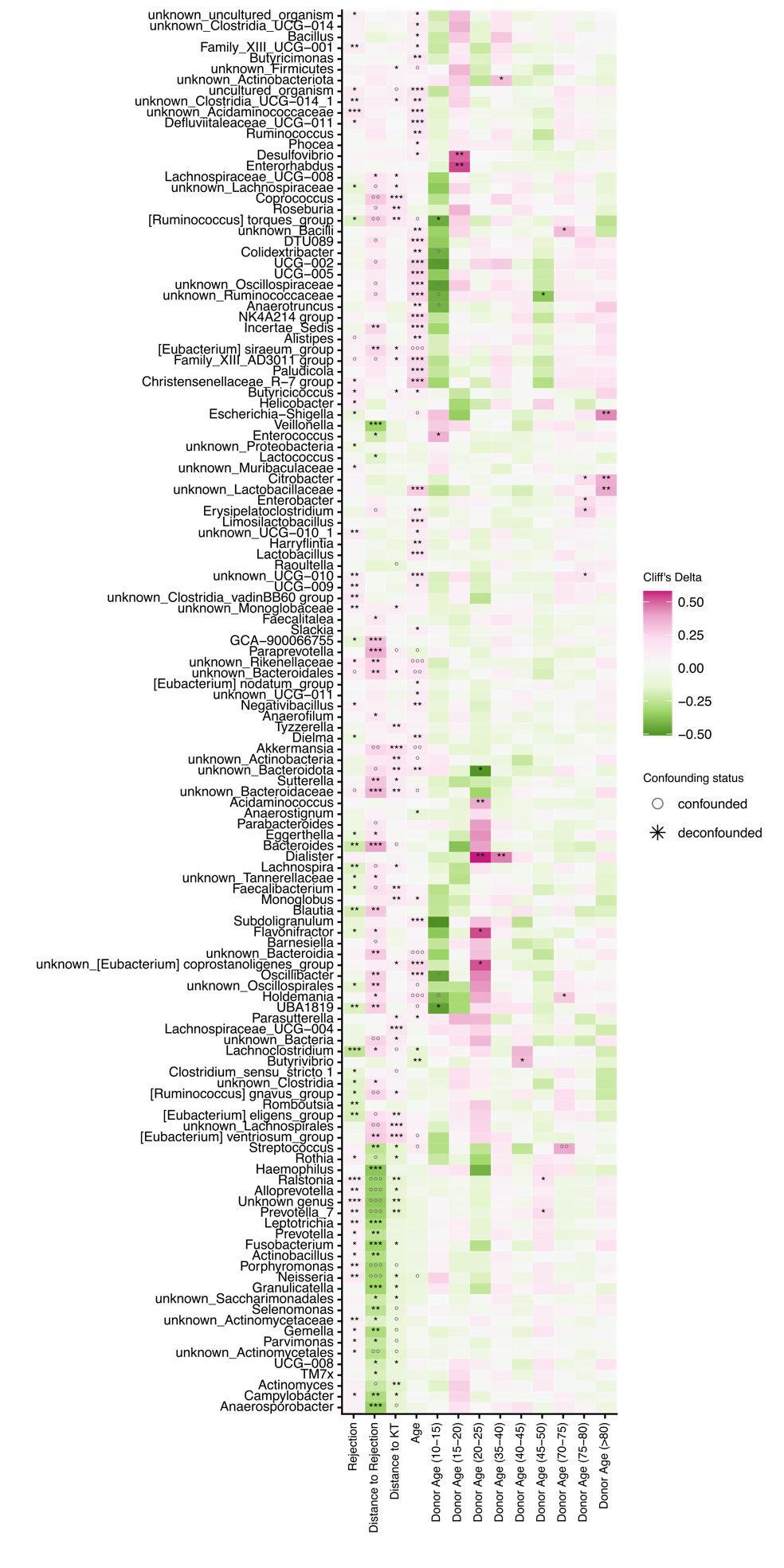

**Supplemental Figure 3. Impact of rejection state on gut microbiome composition.** Analysis of fecal samples from patients experiencing kidney rejection at any time compared to patients never experiencing graft rejection. Heatmap showing associations of bacterial genera with kidney transplant rejection. Meta-variables with significant association are shown. Stars indicate de-confounded significant associations; grey dots indicate confounded associations. °/*FDR< 0.1, °°/**FDR< 0.01, °°°/***FDR< 0.001.

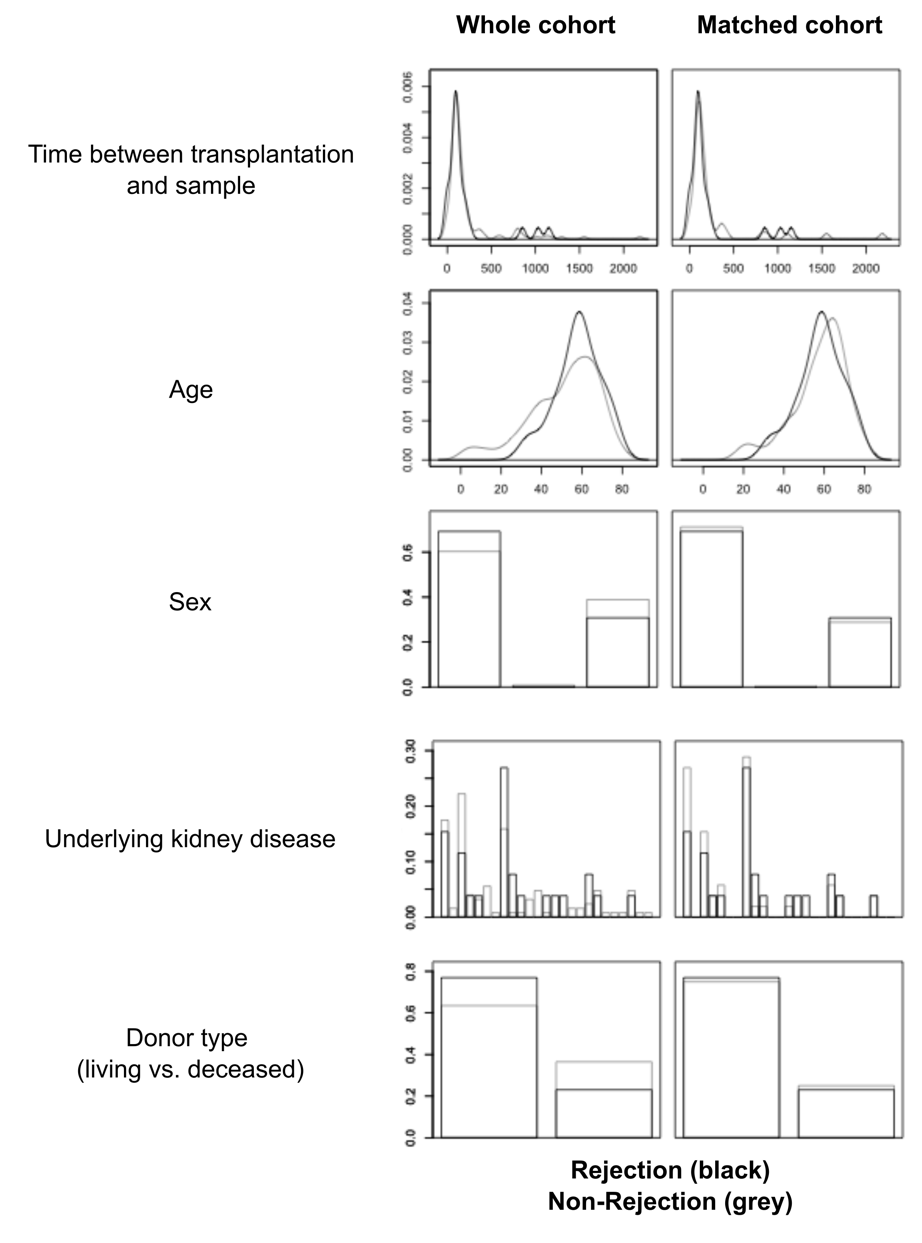

**Supplemental Figure 4: Matching of rejection and non-rejection patients.** Plots indicate metavariables before (left) and after propensity score matching (right). Grey bars/ histogram indicate non-rejection before (left) and after (right) propensity score matching. Rejection is shown in black.

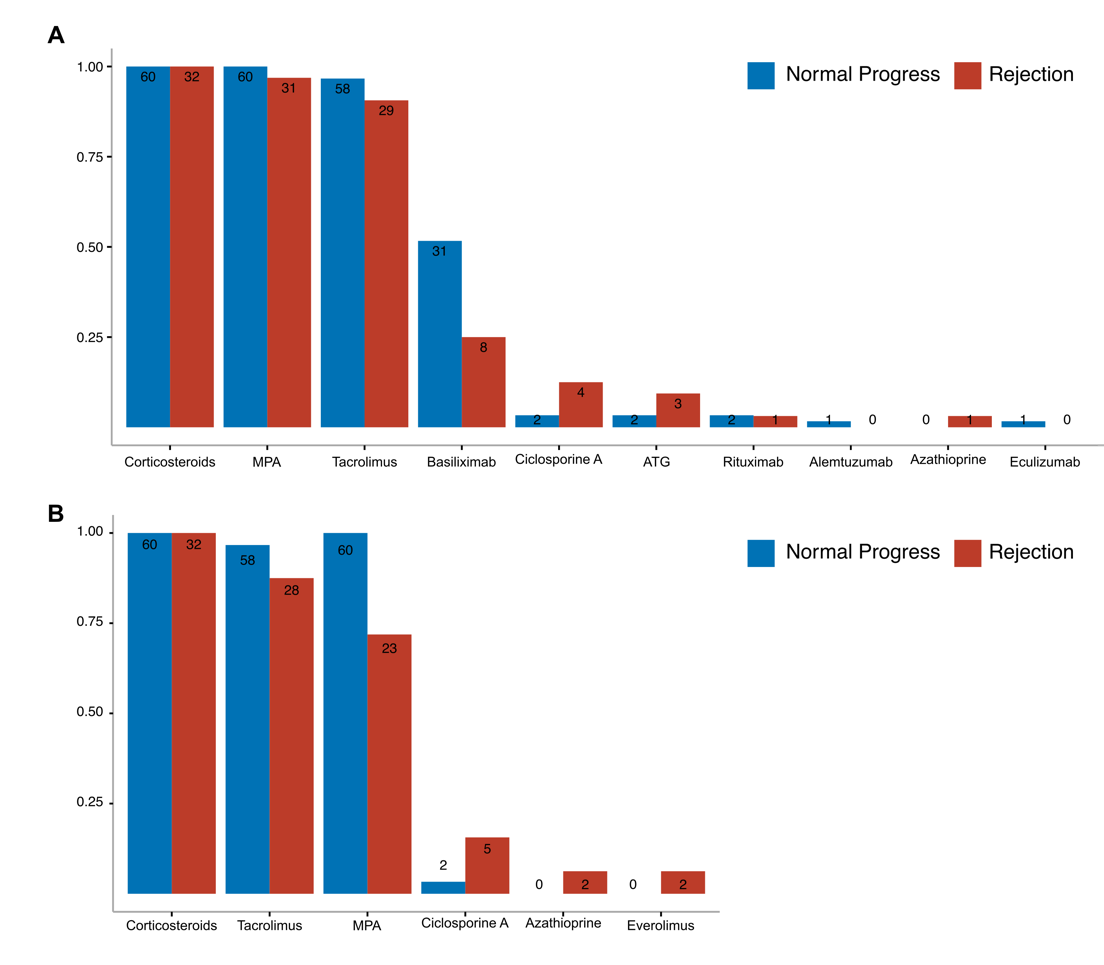

**Supplemental Figure 5: Immunosuppressive treatment at the time of transplantation (A) and at the time of fecal sampling (B).** Fecal samples were analysed from patients before experiencing graft rejection (Rejection, red) and propensity score matched (1:2) controls (normal progress, blue). Bar plots indicate medication at A) transplantation and B) fecal sampling.

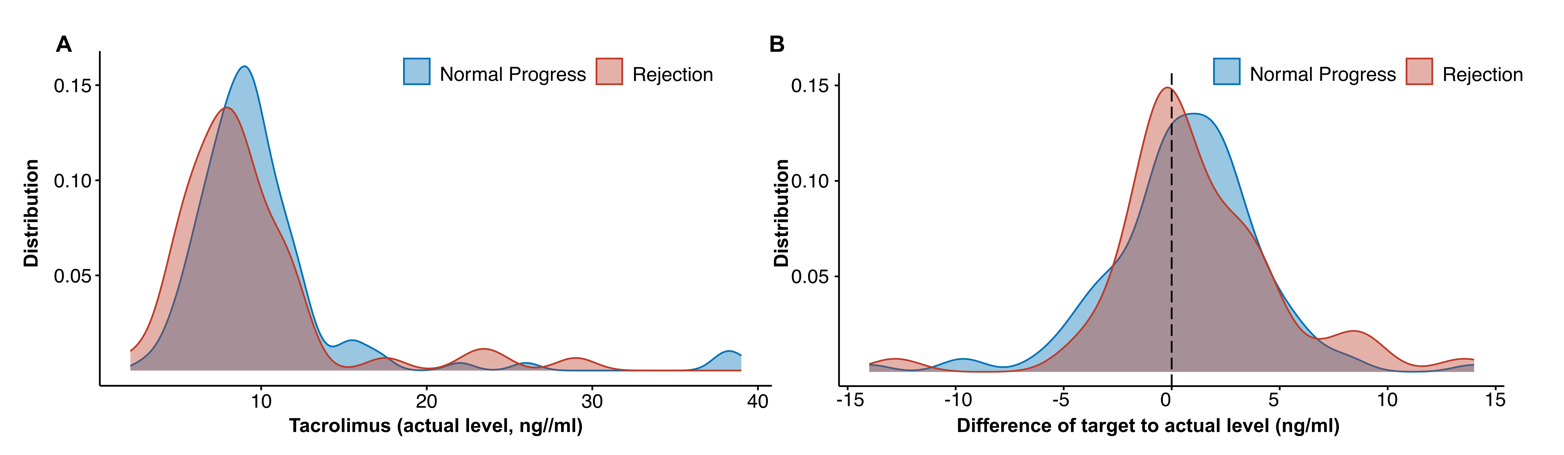

**Supplemental Figure 6. Tacrolimus plasma levels in matched rejection and non-rejection patients.** A) Tacrolimus plasma levels of n=92 patients (matched cohort) for the time between transplantation and sample collection before rejection event (n=121 data points of 51 patienrts for non-rejection and 51 data points of 23 patients for rejection group). B) Difference of target to actual values of Tacrolimus. Target values have been determined by the respective clinical recommendations after kidney transplantation.

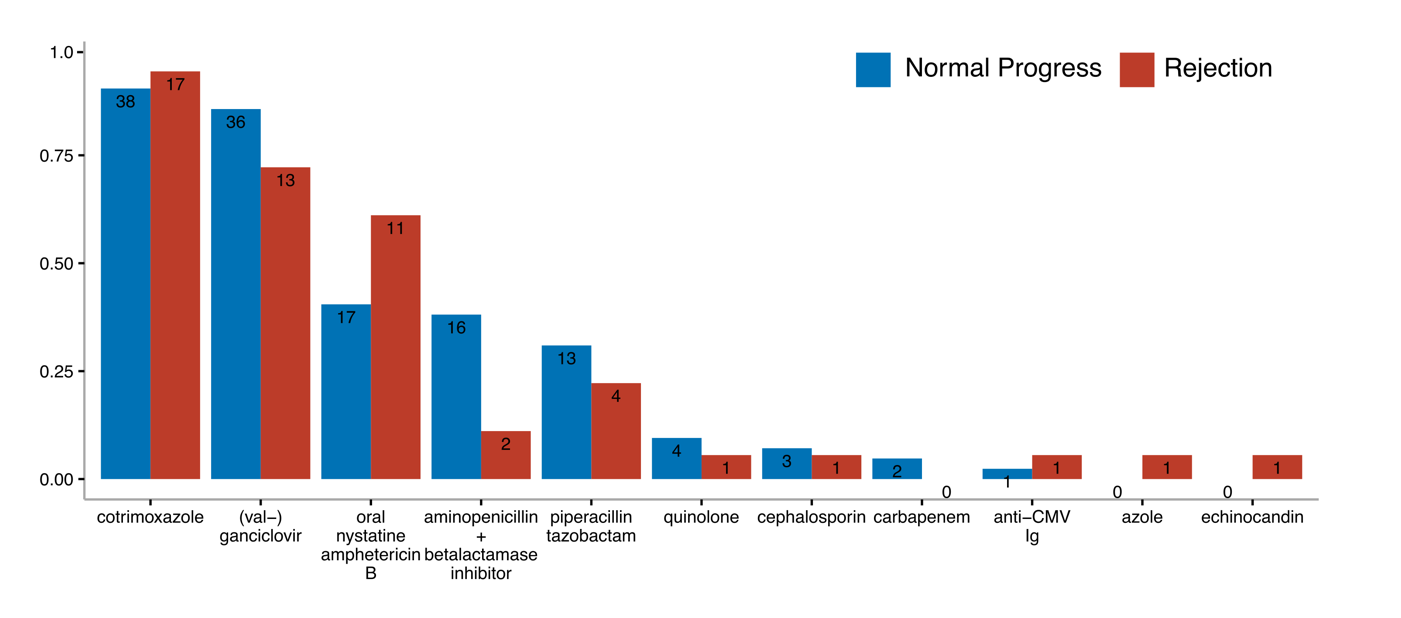

**Supplemental Figure 7: Anti-infective medications used between kidney transplantation and fecal sampling.** Anti-infective medication used between kidney transplantation and fecal sampling for the matched cohort (normal progress, blue; rejection, red). Medication used both preventive and therapeutic is shown.

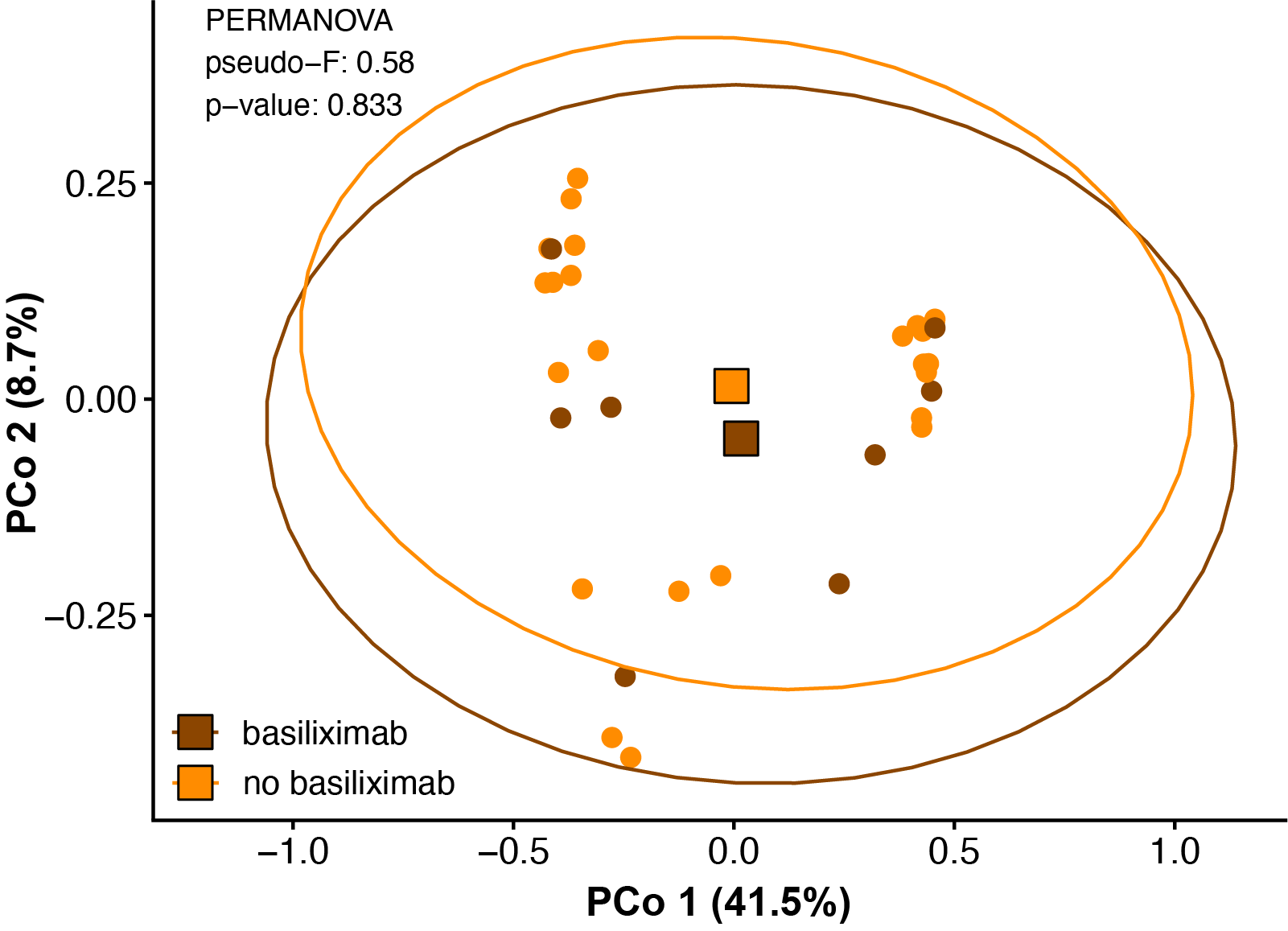

**Supplemental Figure 8: Impact of basiliximab induction therapy on the gut microbiome composition.** PCoA on Bray-Curtis Dissimilarity of the 16S amplicon sequencing in rejection patients with (brown) and without (orange) basiliximab use during induction. Statistical testing using PERMANOVA shows no difference.

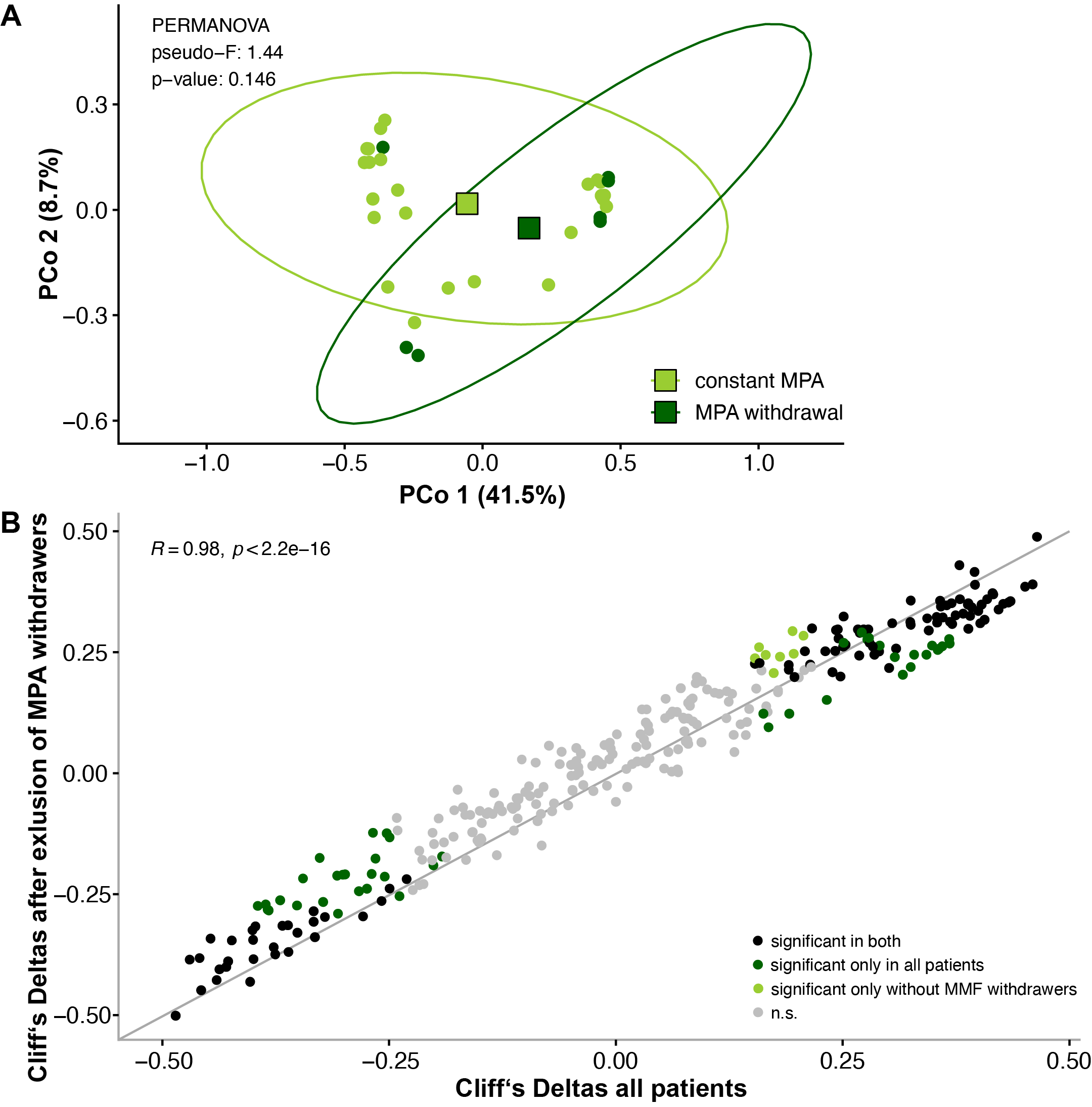

**Supplemental Figure 9: Impact of mycophenole (MPA) withdrawal on the gut microbiome composition.** A) PCoA on Bray-Curtis Dissimilarity of the 16S amplicon sequencing in rejection patients with (light green) and without (dark green) MPA at fecal sampling. Statistical testing using PERMANOVA shows no difference, although some distinct clustering can be seen. B) Correlation of effect sizes in the whole rejection cohort (x-axis) and the cohort excluding patients with MPA withdrawing (y-axis). Axis shows Cliff’s deltas for all detected genera in the gut microbiome. Spearman correlations, individual significances for genera using univariate testing in metadeconfoundR.

**Supplemental Tables**

**Supplemental Table 1:** Clinical variables used for deconfounding of microbiome features

| Variable | Explanation |
| --- | --- |
| Age | Age of the recipient |
| Sex | Sex of the recipient and donor |
| Age donor | Donor age given in 5 years ranges (yes or no) |
| Primary kidney disease | Underlying primary condition in total seven categories |
| Primary kidney disease (ICD code) | Underlying primary condition according to ICD code |
| Type of KT | Deceased or living donation |
| BMI | Body Mass Index of the recipient and donor |
| Height | Body height of recipient and donor in cm |
| Weight | Body weight of the recipient and donor on day of KT in kg |
| Blood group | Blood type of the recipient: A, B, AB, or 0 |
| EBV and CMV serology (IgG and IgM) | Serology status of EBV and CMV before KT for recipient and donor (negative or positive) |
| Viral infection before sample | Any viral infection diagnosed in the time span of KT to sample |
| Viral infection 30d before sample | Any viral infection diagnosed within 30 days before the sample was taken |
| Oral antibiotics before sample | Taken antibiotic in the time span of KT to sample |
| Oral antibiotics 30d before sample | Taken antibiotic within 30 days before the sample was taken |
| Parenteral antibiotics before sample | Any antibiotic given parenteral in the time span of KT to sample |
| Parenteral antibiotics 30d before sample | Any antibiotic given parenteral within 30 days before the sample was taken |
| Number of antibiotic treatments | Total number of antibiotics taken in the time span of KT to sample |
| Number of antibiotic treatments 30d before sample | Total number of antibiotics taken within 30 days before the sample was taken |
| Antibiotic classes | Number of different types of antibiotics taken in the time span of KT to sample |
| Antibiotic classes 30d before sample | Number of different types of antibiotics taken within 30 days before the sample was taken |
| Min. creeatinine | The minimum creatinine value of the patient after KT |
| Albumin | Albumin blood level in g/L |
| CrP | C-reactive Protein blood level in mg/L |
| Creatinine | Creatinine blood level in mg/dL |
| HbA1c | HbA1c blood level in mmol/mol |
| Hemoglobin | Hemoglobin blood level in g/dL |
| Leukocytes | Leukocytes blood level in 1/mul |
| Lymphocytes | Lymphocytes blood level in 1/μL |
| Monocytes | Monocytes blood level in 1/μL |
| Neutrophil | Neutrocytes blood level in 1/μL |
| Phosphate | Phosphate blood level in in mg/dL |
| PTH | Parathyroid hormone(PTH) blood level in pg/mL |
| Urea | Urea level in mg/dL |
| Uric Acid | Uric acid levels in mg/dL |
| Urine Erythrocytes | Lymphocytes urine level in 1/μL |
| Urine Leukocytes | Leukocytes urine level in 1/μL |
| Urine Nitrite | Indication of whether Nitrite was measured (negative or positive) |
| Urine Protein | Protein urine level in g/L |

**Supplemental Table 2:** Primers used for 16S sequencing.

| **Name** | **Sequence 5’ > 3’** |
| --- | --- |
| 341F-ovh | TCGTCGGCAGCGTCAGATGTGTATAAGAGACAG CCTACGGGNGGCWGCAG |
| 785R-ovh | GTCTCGTGGGCTCGGAGATGTGTATAAGAGACAG GACTACHVGGGTATCTAATCC |

Underlined portions indicate the Illumina-specific adapter overhang sequences (16S Metagenomic Sequencing Library Preparation, Illumina).

**Supplemental Table 3:** Bacterial species included to the gene targeting assays for butyrate and propionate synthesis and corresponding primer sequences.

| **Gene** | **Primers** | **Primer sequence** | **Species** |
| --- | --- | --- | --- |
| ***bcd*** | 95F | CGGCTACACCCGTGACTA | *Roseburia Intestinalis* |
|  | 96R | ACCGGAAATGACCATCATCTG | *Faecalibacterium Prausnitzii* |
|  |  |  | *Eubacterium Rectales* |
|  |  |  | *Eubacterium Halli* |
|  |  |  | *Anaerostipes* |
| ***but*** | 548 F | GGMGACTGGGTSGATTAC | *Flavonifactor plautii* |
|  | 549R  550R | TCCACATACATCTCGGTGTG  TAGATATGCATCCGAGCAGAG | *Faecalibacterium Prausnitzii*  *Coproccocus eutactis* |
| ***mmdA* clone 1** | 645F | GTTTCTGCGATGCGTTCAATA | *Bacteriodes vulgatus* |
|  | 650R | CGGAAGGAATCCCGGTACAT | *Bacteriodes ovatus* |
|  |  |  | *Bacteriodes theta* |
|  |  |  | *Bacteriodes fragilis* |
| ***mmdA* clone 2** | 646F | GGAGAAATCCTCGCCAAGTT | *Faecalibacterium prausnitzii* |
|  | 651R | CAGCCTCGCCATTCTGATAA | *Clostridium spp.* |
| **16S** | 19F | ACTCCTACGGGAGGCAGCAGT |  |
|  | 20R | GTATTACCGCGGCTGCTGGCAC |  |

**References**

1.	Karch A, Schindler D, Kuhn-Steven A, et al. The transplant cohort of the German center for infection research (DZIF Tx-Cohort): study design and baseline characteristics. *Eur J Epidemiol*. Feb 2021;36(2):233-241. doi:10.1007/s10654-020-00715-3

2.	Klindworth A, Pruesse E, Schweer T, et al. Evaluation of general 16S ribosomal RNA gene PCR primers for classical and next-generation sequencing-based diversity studies. *Nucleic Acids Res*. Jan 7 2013;41(1):e1. doi:10.1093/nar/gks808

3.	Ren Z, Fan Y, Li A, et al. Alterations of the Human Gut Microbiome in Chronic Kidney Disease. *Adv Sci (Weinh)*. Oct 2020;7(20):2001936. doi:10.1002/advs.202001936

4.	Özkurt E, Fritscher J, Soranzo N, et al. LotuS2: an ultrafast and highly accurate tool for amplicon sequencing analysis. *Microbiome*. 2022/10/19 2022;10(1):176. doi:10.1186/s40168-022-01365-1

5.	Quast C, Pruesse E, Yilmaz P, et al. The SILVA ribosomal RNA gene database project: improved data processing and web-based tools. *Nucleic Acids Research*. 2012;41(D1):D590-D596. doi:10.1093/nar/gks1219
